# Investigating the impact of less than or greater than 60 seconds of inter-set rest on muscle hypertrophy and strength increases in males with >1 year of resistance training experience: systematic review with meta-analysis

**DOI:** 10.1101/2025.09.22.25336351

**Authors:** L. Davidson, S. Barillas

## Abstract

**Background:** Inter-set rest intervals (ISR) influence resistance training adaptations. Shorter intervals (<60s) promote metabolic stress, while longer intervals (>60s) are thought to enhance recovery, mechanical tension, and strength.

**Aim:** The systematic review and meta-analysis examined the effects of <60s vs. >60s ISR on hypertrophy, strength, and secondary outcomes in resistance-trained males.

**Methods:** Six studies met the inclusion criteria, evaluating muscle hypertrophy, strength, metabolic hormones, power output, and motor unit recruitment. Standardised mean differences (SMDs) with 95% confidence intervals (CIs) were calculated, and forest plots were generated to visualise pooled effects.

**Results:** Strength was modestly improved with longer ISRs (SMD = −0.74) but only trivial differences in hypertrophy (SMD = 0.08). Metabolic hormone responses showed negligible variation between conditions (SMD = 0.11). Secondary outcomes were mixed, with motor unit recruitment slightly favouring shorter ISRs (SMD = −0.66), while power output tended to favour longer ISRs (SMD = −0.64). Forest plots illustrated the heterogeneity of effects across studies.

**Conclusion:** Shorter ISRs appear to slightly reduce strength and power outcomes, while longer ISRs offer minimal benefit for hypertrophy. These findings suggest that longer ISRs may not confer superior benefits for muscle growth, as previously assumed in the literature (e.g., Schoenfeld, 2016). The variability across outcomes underscores the need for further research to refine ISR recommendations, particularly in trained individuals.

**What is already known on this topic:**

- Inter-set rest intervals (ISR) influence hypertrophy and strength outcomes in resistance training.
- Short rest (<60 s) increases metabolic stress but may impair recovery and force output.
- Longer rest (>60 s) consistently supports strength gains, but evidence for hypertrophy advantages remains inconclusive.

**What this study adds:**

- Synthesises evidence specifically in resistance-trained males (≥1 year RT experience).
- Shows hypertrophy outcomes are trivial and do not favour longer ISRs over shorter ISRs.
- Confirms longer ISRs modestly improve strength and power, while hormonal and motor unit responses remain inconsistent.
- Underscores the absence of robust evidence supporting longer ISRs as a superior hypertrophy strategy in trained populations.

## Introduction

Resistance training (RT) is a proven method to enhance muscle hypertrophy and strength.^37^ Muscle hypertrophy refers to the enlargement of muscle fibres, primarily through an increase in their cross-sectional area. Consistent resistance training promotes muscle hypertrophy through three primary mechanisms: mechanical tension, muscle damage, and metabolic stress. Mechanical tension, resulting from force generation and stretch during exercise, initiates mechanotransduction pathways that stimulate muscle growth. Muscle damage, characterised by microtrauma to muscle fibers, triggers inflammatory responses and satellite cell activation, facilitating repair and growth. Metabolic stress, arising from the accumulation of exercise-induced metabolites, contributes to hypertrophy by inducing hormonal responses and cellular swelling.^37,39^ Extensive research has shown that regular RT leads to substantial increases in muscle size, especially when paired with effective nutritional and recovery strategies.^44^ Although several reviews have explored hypertrophic adaptations from resistance training and the impact of <60S and >60s inter-set rest (ISR), none have exclusively compared ISR intervals in resistance-trained males, nor have they explored secondary outcomes that may affect hypertrophy and strength gains.

A central factor in determining RT outcomes is intensity, commonly defined as the weight lifted relative to an individual’s one-repetition maximum (1RM).^15^ Higher intensities are associated with greater strength gains, primarily due to the increased mechanical tension applied to muscle fibres.^37^ Mechanical tension creates structural changes in the muscle fibres, stimulating muscle protein synthesis, which is critical for both hypertrophy and strength. This is particularly relevant for experienced lifters, where progressive overload is crucial for the ongoing strength development. Additionally, training volume, the total number of sets and repetitions performed, is another key driver of hypertrophy.^40^ Higher volumes, particularly through multiple sets per muscle group, have been linked to more significant muscle growth. A higher volume increases time under tension and cumulative metabolic stress, both of which are necessary to maximise hypertrophic responses. However, elevated volumes can also induce fatigue, requiring adjustments in variables, such as rest intervals, to ensure adequate recovery and sustained performance.^26^ Initially, research on resistance training focused largely on intensity and volume as primary drivers of hypertrophy and strength.^15,37^ Over time, however, it became clear that other factors, such as rest intervals between sets, played a vital role in optimising these adaptations. The rest intervals were initially viewed as secondary because the focus was on the immediate mechanical tension generated through intensity and volume. However, it has become evident that rest intervals are key in managing fatigue, recovery, and subsequent performance, making them a critical factor in long-term adaptation.^24^ As the understanding of RT has evolved, the importance of rest intervals has become more prominent, particularly for enhancing hypertrophy and strength.^38^

Training outcomes in resistance training are influenced by several modifiable variables, commonly summarised by the acronym CO-FIVR-P (Choice, Order, Frequency, Intensity, Volume, Rest, and Progression). Despite the significance of these variables, rest intervals, while critical, have historically received less attention than intensity and volume, especially regarding their specific impact on hypertrophy and strength in trained individuals.^14^ Rest intervals regulate the effective recovery of muscles between sets, which directly affects subsequent muscle activation, the extent of metabolic stress, and the overall hormonal environment for growth. These rest periods affect recovery, muscle activation, and metabolic stress, all of which contribute to the long-term adaptation. While adjusting intensity and volume can shift the focus between hypertrophy and strength, managing fatigue through rest intervals is equally important in promoting recovery, muscle growth, and performance.^24,34^ Despite considerable research, the optimisation of training variables such as ISR periods remains a crucial area of study, particularly for resistance-trained males.^39,18^ ISR intervals, defined as the time between sets of exercises, significantly influence physiological responses to resistance training.^11^ Shorter rest periods, typically under 60s, are associated with greater metabolic stress, enhancing hypertrophy by increasing muscle fibre recruitment and circulating anabolic hormones.^39^ The heightened metabolic stress results from the rapid accumulation of by-products such as lactate and hydrogen ions, which contribute to the activation of anabolic signalling cascades. One key pathway is the mammalian target of rapamycin (mTOR), a central regulator of muscle protein synthesis and hypertrophy. When these metabolites accumulate beyond a cellular threshold (commonly induced by high-repetition sets and minimal rest intervals) they stimulate upstream regulators that initiate mTOR activation, thereby promoting muscle fibre growth.^9^

It is hypothesised that optimal rest intervals modulate metabolic stress, hormonal responses, and neuromuscular recovery, as central nervous system recovery and peripheral motor unit reactivation are more complete with >60s rest, allowing higher loads and better technique consistency across sets, thereby maximising both hypertrophic and strength adaptations.^5,8^ Conversely, longer rest periods (>60s) are more effective for strength development, as they provide sufficient recovery, allowing higher training volumes and intensities.^20^ Although rest intervals less than 90s are commonly recommended for hypertrophy, there remains an ongoing debate over the optimal duration to maximise the hypertrophic stimulus, particularly at the 60-second mark.^38^ The current review is unique in synthesising recent evidence specifically on inter-set rest intervals in experienced male lifters, filling a critical gap and providing updated, evidence-based recommendations.

The lack of consensus on the ideal rest interval for simultaneously optimising hypertrophy and strength is a recurring issue in literature. Some studies suggest that shorter rest periods can limit strength adaptations due to insufficient recovery,^11^ whereas others argue that longer rest intervals may not induce sufficient metabolic stress for maximal hypertrophy.^1^ This divide often stems from the competing demands of strength training, which requires adequate recovery, and hypertrophy training, which benefits from cumulative metabolic stress over time. These conflicting findings have led to varied recommendations, underscoring the need for evidence-based guidelines to help practitioners balance the rest intervals for both strength and hypertrophy. Research findings on rest intervals have presented complex narratives. For instance, Schoenfeld et al. (2016) found that participants using 180s rest intervals experienced greater muscle hypertrophy over eight weeks compared to those using 60s intervals. In contrast, Gentil et al. (2010) found no significant difference in strength gains between short and long rest periods, though the study used work-to-rest ratios rather than fixed interval lengths. Willardson and Burkett (2008) also highlighted that longer rest intervals (four-minutes) may be more beneficial for strength, but their study’s small sample size and focus on trained male athletes limit the generalisability of these findings. Overall, these mixed results reflect the intricate relationship between rest intervals and RT outcomes, and these discrepancies, including variations in reported effect sizes and differences in study populations, highlight the significant uncertainties in the current literature that the current review aims to resolve

Shorter rest periods increase metabolic stress by accumulating by-products such as lactate and hydrogen ions.^14^ This metabolic environment stimulates growth by activating cellular pathways linked to protein synthesis, such as the mTOR pathway. This stress can drive hypertrophy by promoting greater muscle fiber recruitment and eliciting acute elevations in anabolic hormones such as testosterone, growth hormone (GH), and insulin-like growth factor-1 (IGF-1).^37^ These hormonal responses play pivotal roles in muscle adaptation: testosterone enhances protein synthesis and satellite cell activation; GH stimulates amino acid uptake and collagen synthesis; and IGF-1 mediates muscle cell proliferation and differentiation. Collectively, these hormonal surges contribute to the anabolic environment necessary for muscle growth following resistance exercise.^25,45^ However, shorter intervals have been shown to limit the load lifted in subsequent sets, potentially impairing strength gains.^23^ In contrast, longer rest intervals allow for a more complete recovery, enabling heavier loads and supporting strength development.^42^ Although longer intervals are primarily linked to strength improvements, their role in hypertrophy is less clear, with some suggesting that they may still support hypertrophy through higher total training volumes.^46^

Evidence from recent studies presents a complex and variable picture regarding the role of rest intervals, with evidence supporting both short and long rest durations depending on specific training goals. However, inconsistencies in study designs, participant characteristics, and training backgrounds complicate the interpretation of these findings. A major limitation of existing research is the frequent use of untrained or mixed populations, which may not accurately represent the needs of well-trained individuals with different recovery and performance capacities.^18^ Additionally, the lack of long-term studies on rest intervals leaves unanswered questions regarding their effects over extended training periods. This gap in long-term research limits current understanding of how rest intervals affect strength and hypertrophy across various phases of training. Physiologically, metabolic stress plays a central role in the hypertrophic response to RT,^9^ influenced by the duration of rest intervals. Shorter intervals of less-than 60s, lead to the accumulation of metabolic by-products, such as lactate and hydrogen ions,^14^ which contribute to hypertrophy by activating pathways involved in muscle growth, such as the mTOR pathway.^9^ Although beneficial for muscle growth, short intervals may compromise performance owing to increased fatigue, requiring careful balancing to maximise both hypertrophy and strength.^23^

In addition to metabolic stress, rest intervals significantly affect hormonal responses during RT, particularly the release of anabolic hormones, such as testosterone, growth hormone (GH), and insulin-like growth factor 1 (IGF-1).^25^ Shorter intervals are associated with greater acute hormonal response, which promotes muscle repair and growth (Frystyck, 2010). However, longer intervals allow for heavier loads and strength development despite producing a less pronounced hormonal response.^27^ The effectiveness of longer rest periods can be attributed to their role in facilitating more complete neuromuscular recovery, particularly through the replenishment of adenosine triphosphate (ATP) and phosphocreatine (PCr) stores. These high-energy phosphates are rapidly depleted during high-intensity resistance training and typically require approximately 2–3 minutes for substantial resynthesis. This recovery window is essential for restoring muscular force output in subsequent sets, thereby supporting higher training intensities and improving strength performance.^30,36^

These factors are critical for maintaining the mechanical tension required to stimulate strength adaptation during the subsequent sets.^37^ While shorter rest periods are associated with transient increases in anabolic hormones such as growth hormone, evidence suggests that these acute hormonal responses have a limited direct influence on long-term strength gains.^38^ In contrast, the ability of longer rest intervals to sustain high-intensity efforts has been shown to improve 1RM performance and optimise strength-oriented training protocols.^46^ These findings underscore the importance of aligning rest interval durations with specific training objectives, with longer intervals proving particularly beneficial for strength enhancement.

Optimisation of rest intervals appears essential for improving hypertrophy and strength outcomes, based on current evidence. The current review aims to address the lack of consensus and offer evidence-based recommendations on the impact of rest intervals, particularly those less than or greater than 60s, on muscle hypertrophy and strength in resistance-trained males. The systematic review adhered to the PRISMA 2020 guidelines, ensuring a rigorous and transparent approach to data synthesis and interpretation.

Therefore, the objective of the systematic review was to assess the effects of ISR intervals less than or greater than 60s on muscle hypertrophy and strength gains in resistance-trained males, compared to standard rest intervals, evaluating outcomes such as muscle cross-sectional area, muscle thickness, one-repetition maximum strength, power output, and hormonal responses over a training period of at least 8 weeks. The review compares short (<60s) and long (>60s) ISR intervals on hypertrophy and strength in resistance-trained males with ≥1 year of experience, whilst evaluating primary outcomes including changes in muscle size (cross-sectional area or thickness) and strength (1RM or peak force), as well as secondary outcomes such as power output, metabolic hormone levels, and motor unit recruitment. By considering studies with a minimum duration of 8 weeks to capture meaningful adaptations, the review aims to analyse the potential trade-offs between hypertrophy and strength gains when manipulating rest intervals, ultimately providing evidence-based recommendations for optimising rest intervals in resistance training programs for experienced male lifters seeking to maximise both muscle growth and strength development.

## Methods

### Inclusion/Exclusion Criteria

The inclusion and exclusion criteria were designed to ensure that the studies selected for this review specifically examined the impact of ISR intervals on hypertrophy and strength within a population of resistance-trained males. To enhance the validity and reliability of the findings, studies were rigorously assessed against multiple predefined quality criteria, including methodological rigor, study design (e.g., randomised controlled trials), and the clarity of outcome measures.^21^ This approach ensures that only high-quality evidence is synthesised, minimising bias and enhancing the applicability of the conclusions to resistance-trained individuals.

### Search Strategy

The review included randomised controlled trials (RCTs) and controlled trials (CTs) that implemented resistance training (RT) protocols in resistance-trained males, using ISR intervals of either <60S or >60s as a training intervention.

A systematic literature search was conducted by one author and independently reviewed by a second reviewer on October 18, 2024, using the PubMed database. PubMed was selected as the sole database because of its extensive coverage of peer-reviewed biomedical and exercise science literature relevant to the review.^13^ Additionally, only English-language publications were included to ensure complete comprehension and accuracy of the data extraction.

The search strategy employed a Boolean approach using the operators AND, OR, NOT, and the following keywords and MeSH terms:

(“Inter-set rest” OR “rest intervals” OR “rest periods” OR “shorter” OR “longer” OR “>60 seconds” OR “<60 seconds”) AND (“hypertrophy” OR “muscle” OR “growth” OR “CSA” OR “cross-sectional area” OR “muscle thickness”) AND (“resistance-trained males” OR “trained males” OR “athletes” OR “trained”).

### Study Selection Process

Two independent reviewers screened studies in two phases using Rayyan, a systematic review software designed to streamline screening.^33^ In Phase 1, both reviewers screened titles and abstracts, classifying studies as ‘included,’ ‘excluded,’ or ‘maybe’ based on the predefined eligibility criteria (Table 1). Discrepancies between reviewers were resolved through discussion, no conflicts arose between reviewers, as consensus was reached on all study inclusion decisions. In Phase 2, studies that passed the initial screening underwent full-text review to confirm eligibility. Rayyan was used to facilitate title and abstract screening, providing tools such as keyword highlighting, blind screening, and categorisation. However, no further automation tools were used for full-text screening, data extraction, or bias assessment. All reference lists of included articles were manually cross-checked to identify additional relevant studies and eliminate duplicate records.

**Table 1.** Inclusion and Exclusion Criteria for the Current Systematic Review.

| Component | Inclusion | Exclusion |
| --- | --- | --- |
| Population | Males<br>Aged 18-50 years<br>>1 year specifically compared inter-set rest intervals in resistance-trained males with over 1 year of experience RT experience | Untrained individuals<br>Individuals with underlying health conditions |
| Intervention | Comparison of <60 seconds and >60 seconds rest intervals during RT<br>Period of >8 weeks | No comparison of rest intervals during RT |
| Outcome Measures | Measure muscle CSA or muscle thickness using valid techniques | Do not measure muscle CSA or thickness |
| Study Design | Randomised Controlled Trials<br>Controlled Trials | Observational studies<br>Case studies<br>Qualitative studies |
| Language | English | Not English |

### Data Extraction

Data extraction was performed by a single reviewer using a pre-designed data extraction form tailored to the objectives of the review, and a second reviewer verified the extracted data to ensure accuracy and resolve any discrepancies. Where data on outcome measures were unclear or missing, assumptions were not made, and attempts were made to contact the study authors for clarification and additional details. To ensure consistency, the reviewers thoroughly discussed conflicts after [L. Davidson] (reviewer 1) and [S. Barillas] (reviewer 2) completed the screening process. The process followed the approach recommended by Cochrane for systematic reviews, which suggests that single data extraction with verification can be a valid and efficient method when double data extraction is not feasible owing to time or resource constraints.^21,35^ Therefore, extracted data was checked for accuracy by [L. Davidson] & [S. Barillas] and was entered into Microsoft Excel, double-checking this for accuracy. If the information regarding any of the above was unclear, the authors of the reports would have been contacted to provide further details.

Table 2 includes the outcome variables measured in the accepted studies. Where multiple outcomes were reported, the results that provided the most complete information for analysis were selected. If multiple results remained, all available outcomes (without results) were listed and a content expert was asked to independently rank these based on relevance to the review question, and the validity and reliability of the measures used.

**Table 2.** Eligible Outcome Measures, Assessed Variables and Method Of Measurement.

| Outcome Measure | Variable Assessed | Method of Measurement |
| --- | --- | --- |
| Muscle Hypertrophy (Primary) | Muscle cross-sectional area (CSA) or muscle thickness | Ultrasound, MRI, CT scans, DEXA |
| Muscle Strength (Primary) | One-repetition maximum (1RM) or peak force | 1RM testing, Isokinetic dynamometry |
| Metabolic Hormones (Secondary) | Hormones assessed (e.g., testosterone, growth hormone, IGF-1) | Blood tests, enzyme immunoassays |
| Power Output (Secondary) | Peak power or mean power output | Force plates, velocity-based training |
| Motor Unit Recruitment (Secondary) | Motor unit activation | Electromyography (surface/intramuscular) |

For strength and hypertrophy outcomes, muscle CSA and 1RM were prioritised, followed by power measures, metabolic hormones, and motor unit recruitment as they were most relevant to the current research question.

**Table 3.** Collected Study and Intervention Characteristics.

| Section | Description |
| --- | --- |
| The Report | Author, Year, and Source of Publication |
| The Study | Sample Characteristics: Age, sex, resistance training experience |
|  | Study Design: Randomised controlled trial, observational, etc. |
| The Participants | Inclusion and Exclusion Criteria: Health status, resistance training experience, and demographic details |
|  | Health Status: General health, any specific conditions or diseases |
| The Research Design and Features | Sampling Method: Method used to select participants |
|  | Treatment Assignment: How participants were assigned to groups |
|  | Adherence: Compliance with the intervention, any dropouts or deviations |
|  | Non-response: Missing data or lack of study participation at different stages of the study |
|  | Length of Follow-up: Duration of monitoring after the intervention |
| The Intervention | Rest Interval Duration: Time between sets or exercises during training |
|  | Resistance Training Program: Frequency, type of exercises, intensity, volume, and progression protocols |
|  | Other Relevant Training Variables: Any other variables influencing training (e.g. periodisation) |

By following this systematic approach to data extraction, the most relevant and reliable data were included for analysis, enabling robust conclusions on the impact of rest intervals on muscle hypertrophy and strength.

### Quality Assessment

The quality of included studies was assessed using the Cochrane Risk of Bias 2 (RoB 2.0) tool.^43^ The Cochrane RoB 2 tool was selected for its ability to comprehensively assess the risk of bias across multiple domains in randomised trials. Limitations include subjectivity in domains requiring interpretation, which was minimised through standardised criteria.^32^ This framework evaluates the risk of bias across five domains: (1) bias arising from the randomisation process; (2) bias due to deviations from intended interventions; (3) bias due to missing outcome data; (4) bias in the measurement of the outcome; and (5) bias in the selection of the reported result. Each domain was assessed and categorised as low risk, some concerns, or high risk of bias. Overall, the risk-of-bias judgment for each study was determined by assigning the highest risk rating to any of the five domains.

To maintain consistency, a standardised data extraction form was used during the assessment process. Decisions for each domain were supported by direct evidence from the study protocols and supplementary materials where available. The overall risk of bias rating for each study was determined based on domain-specific judgements. Given that the quality assessment was conducted by a single reviewer, structured workflows and predefined criteria within the RoB 2 tool were employed to mitigate potential bias and ensure rigorous evaluation.^16^ Transparency in decision-making was prioritised, and any ambiguities were resolved using supplementary study documentation.

### Effect Measures

For muscle hypertrophy outcomes, changes in muscle cross-sectional area (CSA) or muscle thickness were synthesised using standardised mean difference (SMD, Cohen’s d) along with 95% confidence intervals (CIs). This effect measure was chosen because the included studies employed different measurement techniques, and SMD facilitates direct comparisons across various scales.^8^ For strength outcomes, such as the one-repetition maximum (1RM) or peak force, the mean difference (MD) with 95% CIs was calculated, as these outcomes are typically reported in a uniform unit (e.g., kilograms), allowing for straightforward comparisons between groups.

For secondary outcomes, power output was analysed using SMD with 95% CIs, since the measurement scale was consistent across studies. In contrast, changes in anabolic hormone levels (e.g., testosterone, growth hormone, IGF-1) and motor unit recruitment were synthesised using SMD with 95% CIs due to variability in measurement methods and units^3^. These choices ensure that appropriate effect measures are applied based on the nature of the data, and they allow the synthesised results to be re-expressed in terms of commonly understood metrics where applicable.

Additionally, thresholds for interpreting effect sizes (e.g., negligible, small, moderate and large) were pre-specified based on established criteria in the literature, which helps in determining the clinical relevance of the findings.^8^

Finally, interventions were categorised by grouping studies based on rest interval durations (<60 s vs. >60 s) and the number of intervals compared. Some studies directly compared one shorter and one longer rest period (e.g., Schoenfeld et al., Fink et al.), whereas others examined multiple duration (e.g., Marshall et al., de Salles et al., Nibali et al., and Padilha et al.). Furthermore, studies were grouped by the outcomes they assessed, ensuring a structured framework for evaluating the impact of ISR duration on muscle hypertrophy, strength, metabolic hormone responses, power output, and motor unit recruitment.

### Categorisation of Interventions and Studies Comparisons

Study intervention characteristics were systematically tabulated and coded according to predefined criteria, including rest interval durations and any additional elements such as rest-pause or inter-set stretching.

The interventions were categorised by grouping studies based on their rest interval duration (<60 s vs. >60 s) and the number of intervals compared. Decisions regarding study eligibility for each synthesis were made by comparing these tabulated characteristics with the planned synthesis groups. Some studies (e.g., Schoenfeld et al. and Fink et al.) directly compared one shorter and one longer rest period, whereas others (Marshall et al., de Salles et al., Nibali et al., and Padilha et al.) examined multiple rest durations. In addition, four studies focused exclusively on rest interval variations, whereas others incorporated additional elements, such as the rest-pause method used by Marshall et al. (where an initial set was performed to failure, followed by a 20s rest) and inter-set stretching investigated by Padilha et al. The studies were further grouped by their outcomes: several evaluated both muscle hypertrophy and strength (as in Schoenfeld et al. and Fink et al.), some focused solely on strength (e.g., de Salles et al.), while others examined motor unit recruitment (via EMG in Marshall et al. and Padilha et al.), metabolic hormone responses (Fink et al.), and power output (Nibali et al.). This categorisation was performed using a structured framework designed to reduce subjectivity in grouping studies for synthesis. This structured framework ensured a consistent evaluation of the impact of ISR duration on various outcomes. For studies that included multiple intervention components, categorisation was based on the primary component emphasised by the authors, and these studies were allocated to the appropriate synthesis groups. Any ambiguities in the categorisation process were resolved through discussion and, when necessary, consultation with a content expert.

### Data Preparation for Presentation and Synthesis

Several steps were undertaken to prepare the extracted data to ensure consistency in data presentation and synthesis. Where studies reported standard errors (SE), these were converted to standard deviations (SD) using established formulae from the Cochrane Handbook for Systematic Reviews of Interventions.^21^ All calculations were performed in Microsoft Excel (version 16.93.1) to ensure accuracy and reproducibility. Specifically, within-group standard deviations were derived using the following formula:

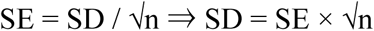

where N_S_ and N_L_ represent the sample sizes of the <60S and >60s ISR groups, respectively. This formula was sourced from Section 6.3 of the Cochrane Handbook for Systematic Reviews of Interventions.^21^ When effect sizes were presented in a format inconsistent with the synthesis requirements (for example, odds ratios instead of standardised mean differences, SMD), the necessary transformations were performed using conversion equations outlined in the literature.^3,8^ All data transformations were applied consistently across the studies, and all underlying assumptions were explicitly stated.

To calculate the standardised mean difference (SMD), commonly known as Cohen’s d, the mean difference between the two groups was first determined. This difference was then divided by the pooled standard deviation (SD_pooled_), which was calculated as follows:

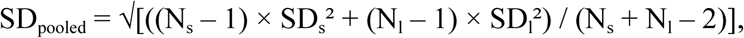

where N_s_ and N_l_ represent the sample sizes of the experimental and control groups, respectively, and SD_s_ and SD_l_ are their corresponding standard deviations. This method standardises the effect by accounting for variability in the data, thus enabling meaningful comparisons across studies with different measurement scales.

For studies that did not report confidence intervals (CIs) for effect measures, CIs were calculated from the available data using standard statistical methods. To derive the 95% CIs for the SMDs, the standard error (SE) of the effect estimate was first calculated using established formulae.^3^ The CI was then computed using the following formula:

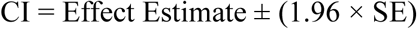

to 1.96, representing the z-value for a 95% CI under a normal distribution. Although a t-distribution may be more appropriate for small sample sizes, the normal approximation was considered sufficiently robust for the analysis.

Since no imputations were made, none were documented. These detailed procedures ensured that the data preparation process was transparent and replicable, allowing for the independent verification of all analyses.

### Presentation of Study Results and Data Visualisation

The results from individual studies were systematically tabulated using a summary table (Table 5), which presents key study characteristics, including sample size, intervention details, and primary and secondary outcome measures. The table was structured by outcome domain and intervention characteristics to facilitate comparisons between studies and to highlight variations in the risk of bias and effect sizes. For example, studies were ordered by descending effect size for muscle hypertrophy, prioritising the most substantial effects.

In addition to the summary table, forest plots were generated to visually represent the effect sizes across studies for the main outcome measures: muscle hypertrophy, muscle strength, and overall outcome measures across studies. These plots provide a clear graphical summary of the pooled standardised mean differences (SMDs) and their associated 95% confidence intervals, assisting in the synthesis of the results and offering a clear visual assessment of the magnitude and direction of the effects. Forest plots also highlight the variability between studies, illustrating the range of effect sizes and the potential impact of individual studies on the pooled results.

Forest plots were included to specifically display the pooled effects for muscle hypertrophy across studies, muscle strength across studies, and all outcome measures across studies. The first plot visualises the pooled SMD for muscle hypertrophy across relevant studies, allowing for a clear comparison of the effects of different inter-set rest durations on muscle growth. The second plot focuses on strength-related outcomes, displaying the pooled effect of inter-set rest intervals on strength gains, as measured by variables such as 1RM and maximal force output. The third forest plot offers a holistic view of the impact of inter-set rest on all relevant outcomes (hypertrophy, strength, metabolic hormones, power output, and motor unit recruitment) across all studies, providing a comprehensive understanding of the effects of inter-set rest intervals in resistance training.

All effect-size conversions and calculations were performed using Microsoft Excel (version 16.93.1). Standard methods for converting standard errors to standard deviations and for calculating pooled SMDs were employed, as outlined in Section 6.3 of the Cochrane Handbook for Systematic Reviews of Interventions^21^ and in meta-analyses texts.^3,8^ All data transformations and adjustments were applied consistently across the studies to ensure transparency and replicability.

These visual and tabulated displays assist in identifying patterns in effect estimates and offer an informal exploration of the variability across studies. The inclusion of forest plots enhances the clarity of data synthesis and supports the overall interpretation of the findings, ensuring that the results are presented transparently. All tables and figures have been consistently referenced throughout the review.

### Methods Used To Synthesise Results

The systematic review examined the effects of inter-set rest intervals (ISR) on muscle hypertrophy, strength, metabolic hormones, power output, and motor unit recruitment. A formal meta-analysis was conducted using pooled standardised mean differences (SMDs) with 95% confidence intervals (CIs) for studies that reported comparable outcomes. This allowed for a quantitative comparison of the effects of <60S versus >60s. Pooled SMD values were derived using Hedges’ g adjustment to account for the small sample sizes.^3^ Forest plots were generated to visually represent the effect sizes for each outcome measure and provide a clear overview of the results across studies.

Given the variability in effect estimates observed across studies,primarily due to differences in study methodologies, participant characteristics, and measurement scales, a formal meta-analysis was conducted to combine the results where appropriate. The forest plots produced illustrate the pooled SMDs for muscle hypertrophy, muscle strength, and overall outcomes, summarising the direction and magnitude of the effects across the studies. However, substantial variability was noted in the SMD values, which was not solely due to random sampling error, but largely due to methodological differences among studies. This variability was captured in the forest plots, which reflects the broader patterns in the data, yet demonstrates the complexity of the synthesis.

All effect size conversions and data synthesis, including the calculations of pooled SMDs and associated CIs, were performed using Microsoft Excel (version 16.93.1), adhering to standard systematic review and meta-analysis methodologies outlined in the Cochrane Handbook for Systematic Reviews of Interventions^21^ and meta-analyses texts.^3,8^

All data transformations and adjustments were applied consistently across the studies to ensure the transparency and replicability of the results.

In summary, the review employed both narrative synthesis and meta-analytic visualisation to synthesise the effects of ISR across studies. The forest plots were integral in representing the range of effect sizes, whereas the pooled SMDs provided a clear quantitative comparison of the effects on key outcomes.

### EDI Statement

This review was conducted in line with BJSM’s Equity, Diversity and Inclusion (EDI) guidelines. The study team considered equity and inclusivity at all stages of the review process, including screening and interpretation. The included studies predominantly featured young, resistance-trained males; therefore, findings may not be generalisable to females, older adults, or populations with limited training experience. This limitation is explicitly acknowledged in the discussion.

## Results

### Search and Selection

The initial database search yielded 1,038 records. After removing duplicates, 1,037 records remained for screening. Following the title and abstract screening process, 1,002 records were excluded, leaving 35 full-text articles for the eligibility assessment. After reviewing the full texts, 29 studies were excluded due to ineligible study design, outcome measures, population, or intervention characteristics, resulting in the inclusion of six studies in the systematic review (Schoenfeld et al., Fink et al., de Salles et al., Nibali et al., Marshall et al., Padilha et al.). No automated screening tools were used in the selection process. A subsequent manual search of the reference lists and citation tracking of the included studies did not yield any additional articles that met the inclusion criteria. Figure 1 shows the PRISMA 2020 flow diagram, which illustrates the number of records at each stage of the selection process.

**Figure 1.** PRISMA 2020 flow diagram showing study identification, screening, eligibility, and inclusion for the systematic review and meta-analysis.

### Excluded Studies

During the full-text screening, 29 studies that initially met the inclusion criteria were excluded. The primary reasons for exclusion were ineligible study designs, inappropriate comparator groups, and insufficient outcome reporting. For example, studies such as Ratamess et al. (2009) and Hartmann et al. (2015) were excluded because of their non-randomised designs, while others (e.g., Scott et al., 2016; Senna et al., 2022) were excluded because they lacked essential outcome data. In some instances, the full-text or critical data necessary to inform eligibility were not accessible, further justifying their exclusion. A comprehensive list of all excluded studies, along with specific reasons for exclusion, is presented in Table 4 (Characteristics of Excluded Studies). Studies cited in italics indicate those excluded for multiple reasons (e.g., failing to meet both study design and population criteria). This detailed documentation allows readers to assess the validity and applicability of review findings.

**Table 4.**
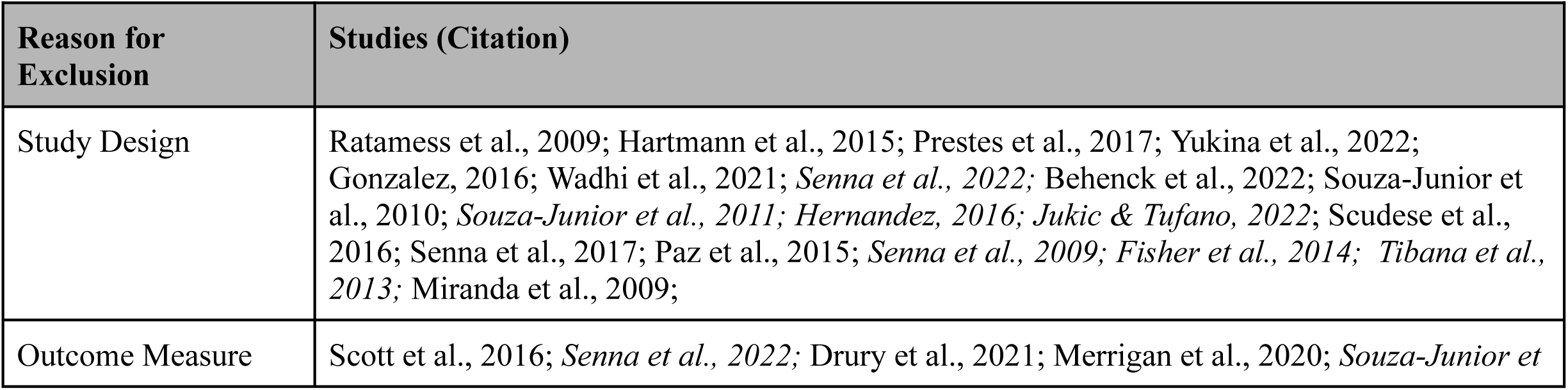
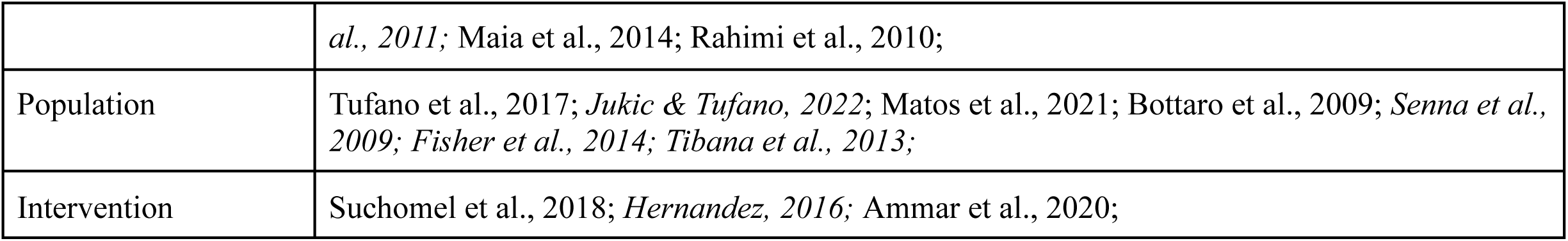
Characteristics of Excluded Studies.

| Reason for Exclusion | Studies (Citation) |
| --- | --- |
| Study Design | Ratamess et al., 2009; Hartmann et al., 2015; Prestes et al., 2017; Yukina et al., 2022; Gonzalez, 2016; Wadhi et al., 2021; *Senna et al., 2022*; Behenck et al., 2022; Souza-Junior et al., 2010; *Souza-Junior et al., 2011*; *Hernandez, 2016*; *Jukic & Tufano, 2022*; Scudese et al., 2016; Senna et al., 2017; Paz et al., 2015; *Senna et al., 2009*; *Fisher et al., 2014*; *Tibana et al., 2013*; Miranda et al., 2009; |
| Outcome Measure | Scott et al., 2016; *Senna et al., 2022*; Drury et al., 2021; Merrigan et al., 2020; *Souza-Junior et* |
|  | *al., 2011*; Maia et al., 2014; Rahimi et al., 2010; |
| Population | Tufano et al., 2017; *Jukic & Tufano, 2022*; Matos et al., 2021; Bottaro et al., 2009; *Senna et al., 2009*; *Fisher et al., 2014*; *Tibana et al., 2013*; |
| Intervention | Suchomel et al., 2018; *Hernandez, 2016*; Ammar et al., 2020; |

In the systematic review examining the effects of ISR durations on muscle hypertrophy, strength, metabolic hormones, power output, and motor unit recruitment, the included studies are summarised in Table 5. Each study is categorised based on its design, participant characteristics, training intervention, and key outcomes, allowing for direct comparisons of how different ISR durations influence training adaptations.

**Table 5.**
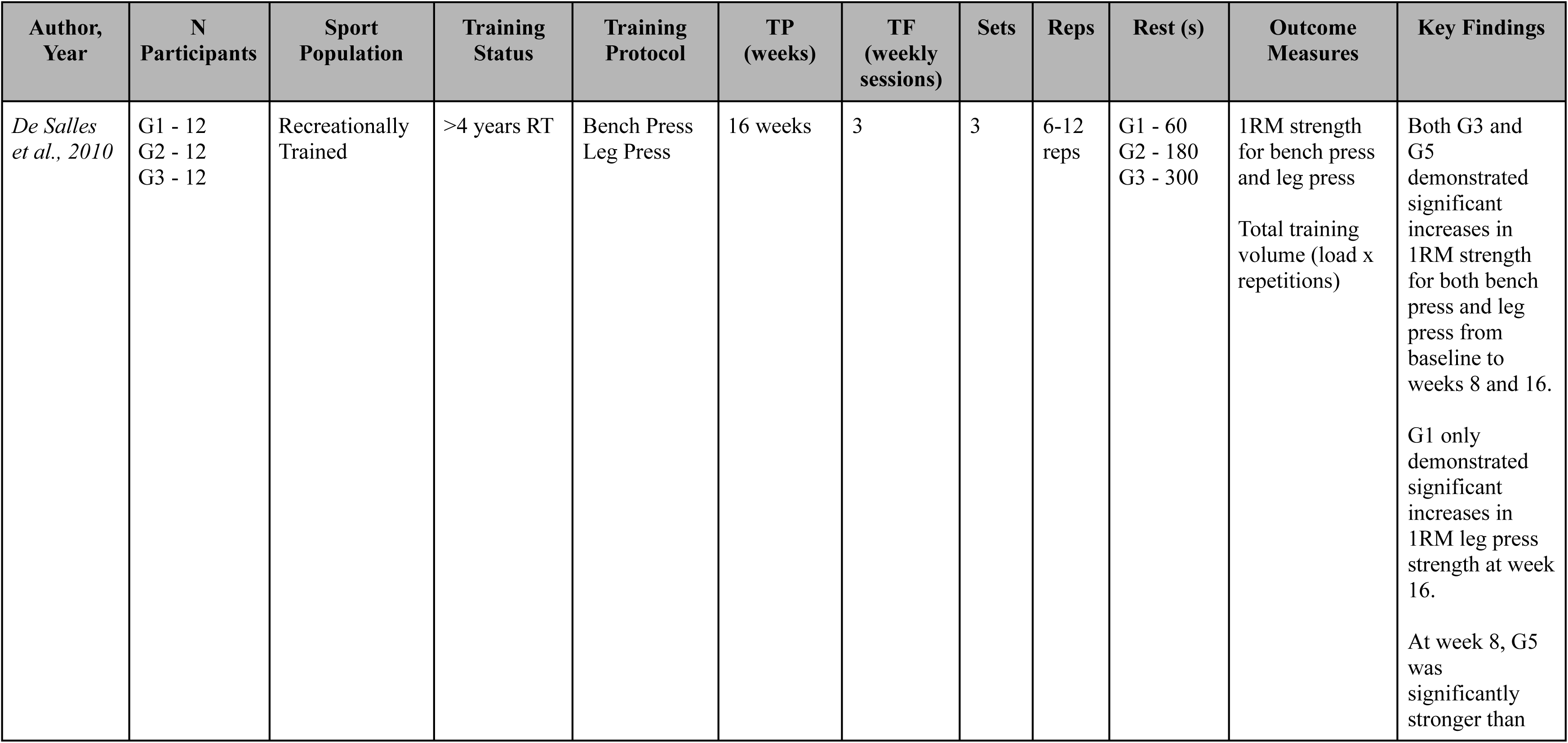
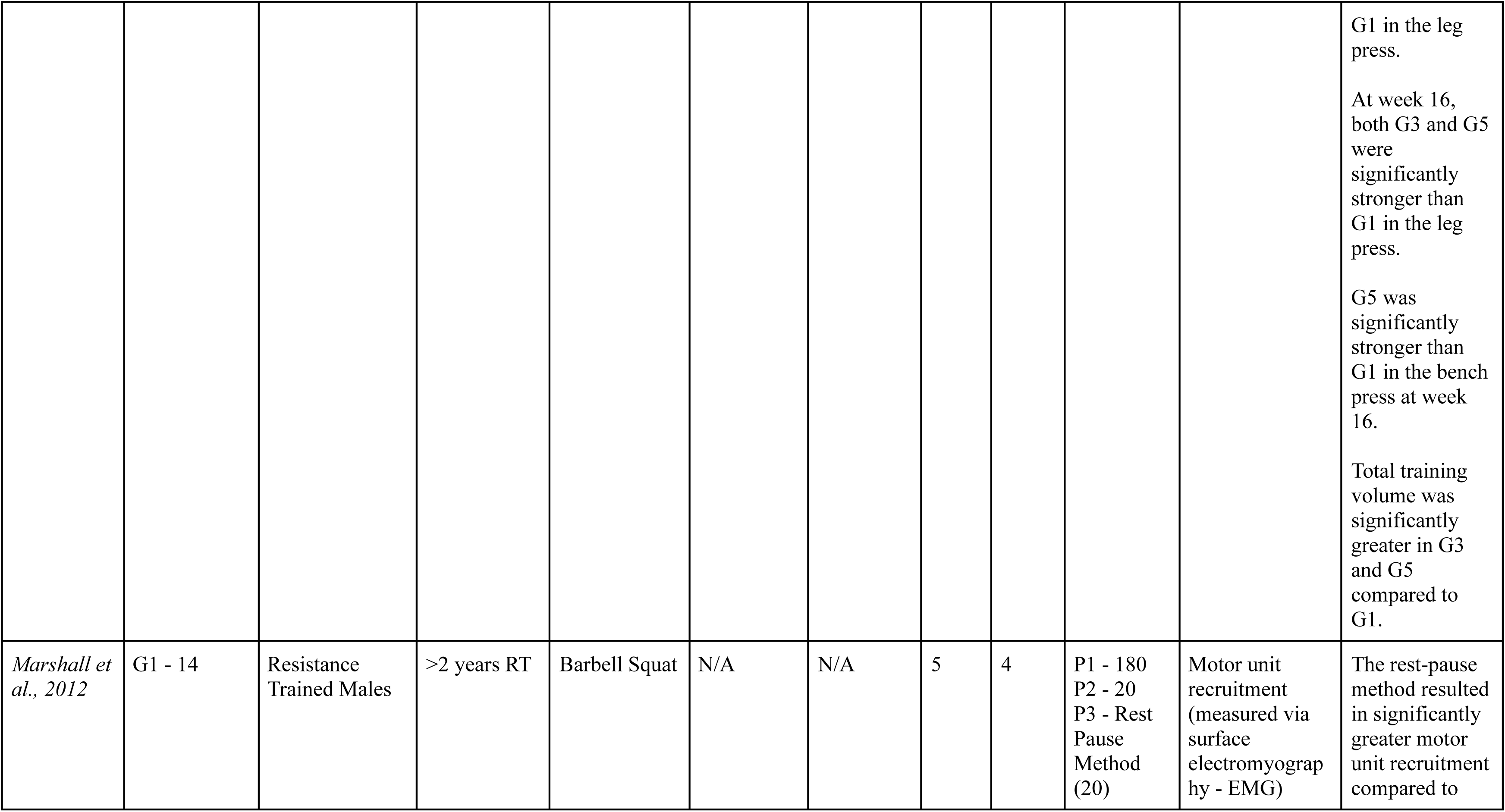
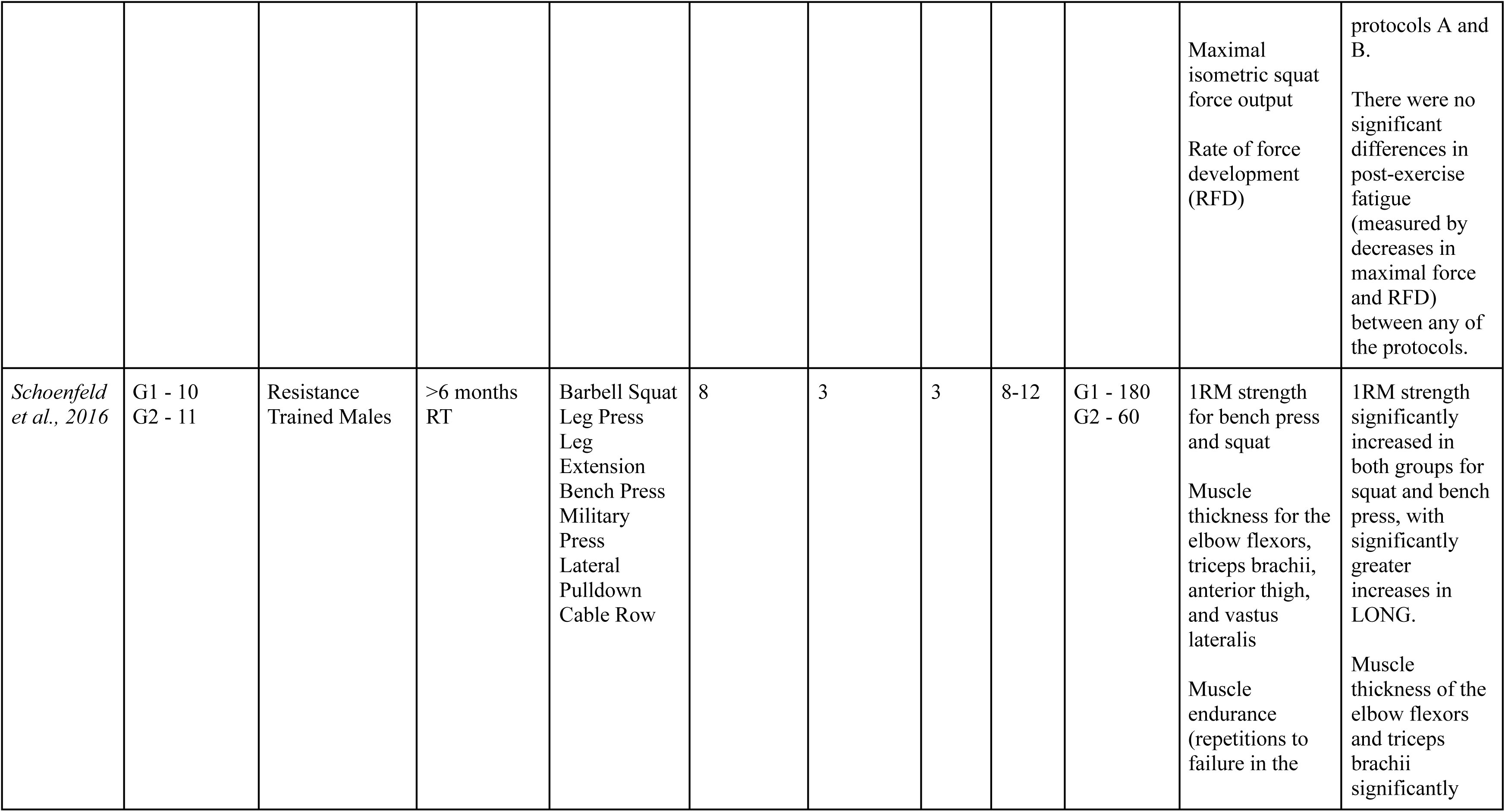
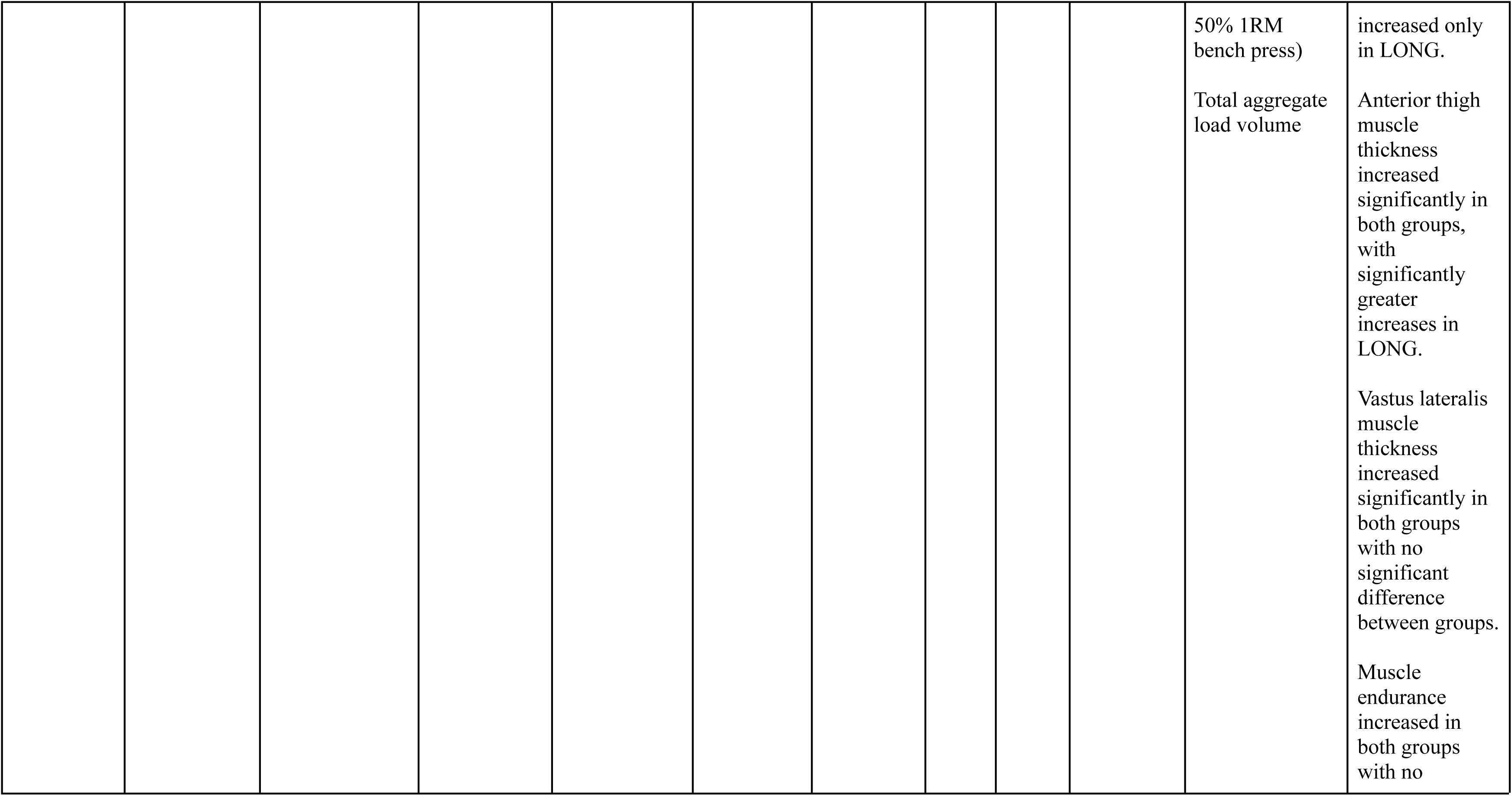
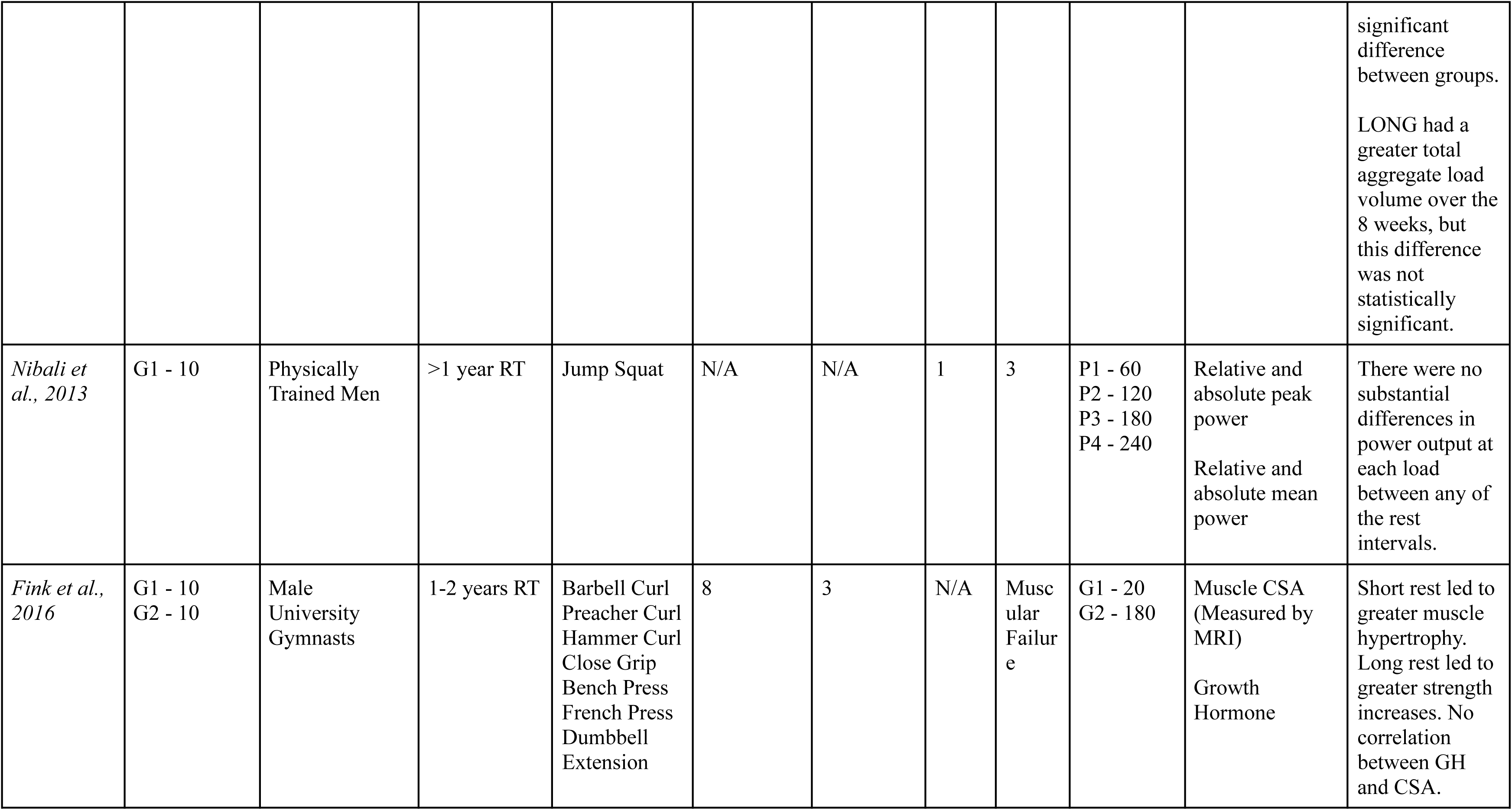
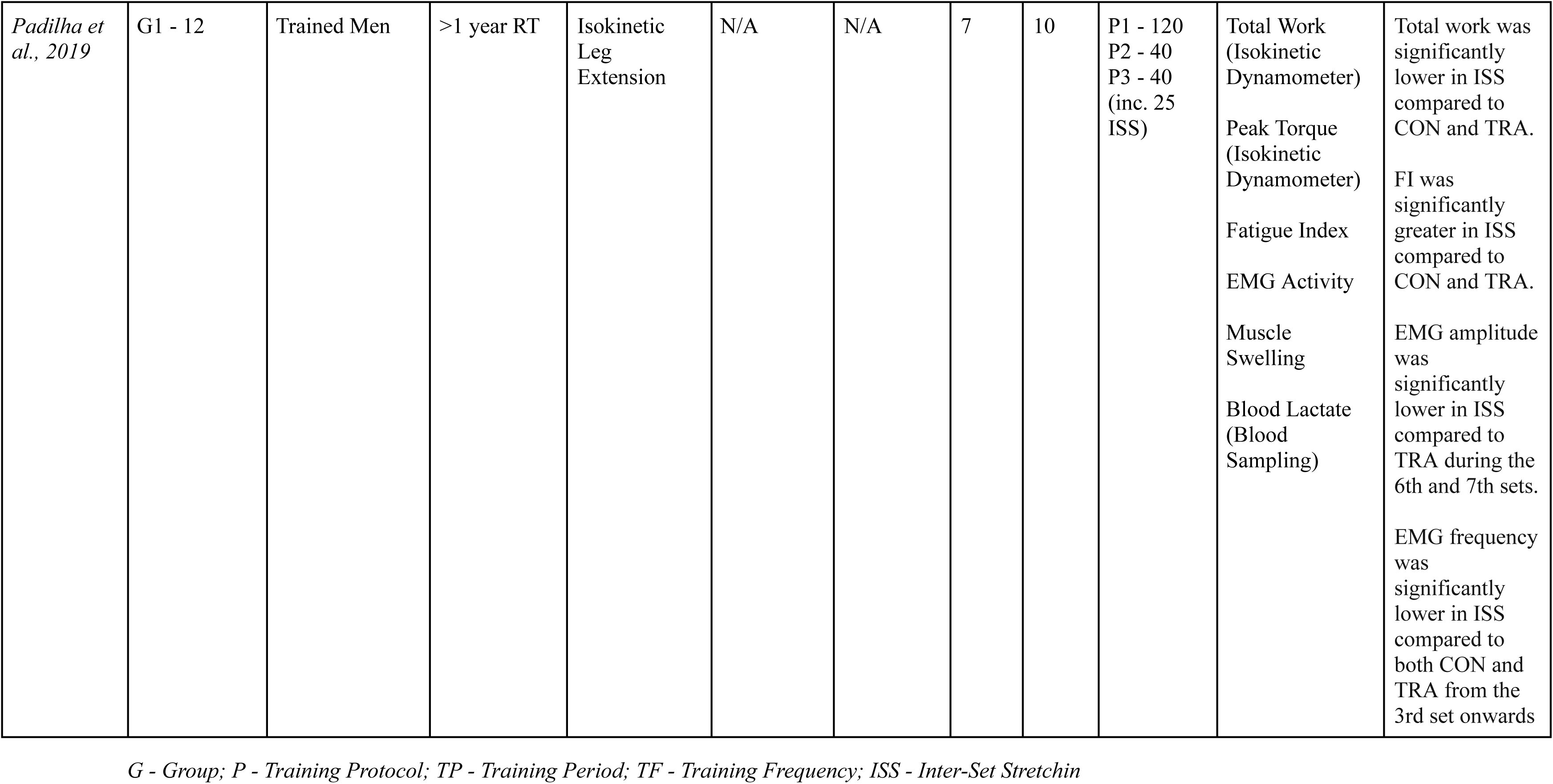
Experimental Details of the Studies Included in the Review.

### Risk of Bias Assessment

The risk of bias was assessed using the Cochrane Risk of Bias 2 (RoB 2.0) tool.^43^ The RoB 2.0 tool was chosen for its comprehensive evaluation of bias across multiple domains in randomised trials, including: (1) bias arising from the randomisation process; (2) bias due to deviations from intended interventions; (3) bias due to missing outcome data; (4) bias in measurement of outcomes; and (5) bias in the selection of the reported result. Each domain was assessed and categorised as low risk, some concerns, or high risk of bias, with the overall risk of bias for each study determined by the highest risk rating among these domains. Justifications for each risk-of-bias judgment (e.g., relevant quotations or specific criteria from the study reports) are provided below Table 6.

**Table 6.** Summary of Results for the RoB 2 Assessment.

| Study | Randomisation Process | Deviations from Intended Interventions | Missing Outcome Data | Measurement of Outcome | Selection of Reported Results | Overall Risk of Bias |
| --- | --- | --- | --- | --- | --- | --- |
| Nibali et al., 2013 | Low | Some Concerns | Low | Low | Low | Low |
| De Salles et al., 2010 | Low | Low | Low | Low | Low | Some Concerns |
| Marshall et al., 2012 | Low | Some Concerns | Low | Low | Some Concerns | Some Concerns |
| Fink et al., 2016 | Low | Some Concerns | Low | Some Concerns | Low | Some Concerns |
| Padilha et al., 2019 | Low | Some Concerns | Low | Low | Low | Low |
| Schoenfeld et al., 2016 | Low | Some Concerns | Low | Low | Low | Low |

The risk of bias assessment was performed by a single reviewer using a standardised data extraction form. Any ambiguities were resolved by referring to the supplementary study documentation. No modifications were made to the RoB 2.0 tool or the ROBVIS tool. A standardised data extraction form was used during the assessment process, and decisions for each domain were supported by direct evidence from the study protocols and supplementary materials.

The results of these assessments are presented in Table 6, along with visual representations using ‘Traffic Light’ Plots (Figure 2) and Weighted Bar Plots (Figure 3) using the ROBVIS tool^28^. The purpose of these visual displays is to enhance transparency by clearly showing the risk-of-bias ratings for each domain and facilitating the identification of patterns or areas of concern across studies. In cases where risk-of-bias assessments were conducted for specific outcomes, they were integrated into the overall study-level judgment.

**Figure 2.** Risk of bias “traffic light” plots illustrating domain-level judgements for each included study.

**Figure 3.** Weighted bar plots showing the distribution of risk-of-bias judgements across all bias domains.

### Justification of Risk of Bias Scores

Nibali et al. (2013) were rated as “some concerns” in the domain of deviations from intended interventions, as the allocation sequence was not fully concealed, potentially leading to minor deviations in intervention delivery. However, the randomisation process and missing outcome data were robust.

De Salles et al. (2010) raised some concerns regarding deviations from intended interventions, as both participants and interventionists were aware of assignment, which may have affected protocol adherence. Additionally, concerns were noted in the measurement domain, where assessor awareness could have influenced the outcomes.

Marshall et al. (2012) were assigned “some concerns” to the deviations from intended interventions domain due to the unblinded delivery of interventions. Concerns were also raised in the measurement domain as the outcome assessors were not blinded, introducing the potential for bias.

Schoenfeld et al. (2016) had some concerns regarding the deviations from the intended intervention domain, as both participants and staff were aware of the intervention allocation. While the intervention was applied consistently, these concerns contributed to the “some concerns” rating, despite low risk in other domains like randomisation and missing data.

Padilha et al. (2019) were rated “some concerns” for deviations from intended interventions due to a lack of blinding during intervention delivery, which may have influenced performance outcomes in the context of inter-set stretching.

Fink et al. (2016) received ‘some concerns’ ratings for deviations from intended interventions and measurement of outcome, while the remaining domains were assessed as low risk.

### Results of Individual Studies

Results of each outcome are presented by detailing, for each study, the measured outcome (e.g., elbow flexor thickness in centimeters), the corresponding effect estimate expressed as a standardised mean difference (SMD) along with its 95% confidence interval (CI), and an interpretation of the magnitude and direction of the effect (e.g., whether the effect favours longer (>60 s) or shorter (<60 s) inter-set rest intervals). This structured presentation - summarised in Table 7 - enables readers to assess each study’s contribution to the review’s overall findings and supports future re-analysis or updates.

**Table 7.** Results of Individual Studies: Effect Sizes and Confidence Intervals for Each Outcome Measure.

| Study | Measure | SMD | 95% CI |
| --- | --- | --- | --- |
| Schoenfeld et al., 2016 | Elbow flexor thickness (cm) | -0.78 | -1.67, 0.16 |
|  | Triceps brachii thickness (cm) | -0.41 | -1.28, 0.45 |
|  | Anterior quad thickness (cm) | 0.79 | -0.1, 1.68 |
|  | Vastus lateralis (cm) | -0.07 | -0.93, 0.79 |
|  | 1RM bench press (kg) | -0.29 | -1.15, 0.57 |
|  | 1RM back squat (kg) | -0.24 | -1.1, 0.62 |
| Marshall et al., 2012 | Maximal Isometric Force Output | -0.55 | -1.31, 0.2 |
|  | Squat RFD | -0.74 | -1.58, 0.1 |
| Fink et al., 2016 | Muscle Cross Sectional Area | 1.29 | 0.31, 2.26 |
|  | Muscle Strength | 0.65 | -0.25, 1.55 |
|  | GH Concentration | 0.11 | -0.77, 0.99 |
| Nibali et al., 2013 | Relative Peak Power 1-min v 2-mins | 0.07 | -0.81, 0.94 |
|  | Relative Peak Power 1-min v 3-mins | -0.8 | -0.96, -0.8 |
|  | Relative Peak Power 1-min v 4-mins | 0.06 | -0.81, 0.94 |
| Padilha et al., 2019 | EMG Activity | -0.66 | -1.49, 0.16 |
| De Salles et al., 2010 | Training Volume G1 v G3 | -0.55 | -1.31, 0.2 |
|  | Training Volume G1 v G5 | -3.93 | -5.34, -2.52 |

*Note: A negative SMD favours longer ISR (>60s), while a positive SMD favours shorter ISR (<60s).*

### Synthesis of Results

The review synthesised the effects of inter-set rest intervals (ISR) on muscle hypertrophy, strength, metabolic hormones, power output, and motor unit recruitment through both meta-analysis and narrative synthesis. Pooled standardised mean differences (SMDs) with 95% confidence intervals (CIs) were calculated for studies reporting comparable outcome measures, allowing for a quantitative comparison of the effects of <60S versus >60s ISR, with SMD values adjusted using Hedges’ g to account for small sample sizes^3^. Forest plots were generated for each outcome measure to visually display the data, illustrating the effect sizes and variability across the studies.

The meta-analysis revealed the overall direction and magnitude of the effects, with the forest plots providing a clear visual representation. In the analysis, negative SMD values indicate a greater effect in favour of longer ISR (>60s), while positive SMD values reflect an effect in favour of shorter ISR (<60s). This orientation was applied consistently across all outcomes While the pooled SMDs for hypertrophy and strength were moderate to large, secondary outcomes such as power output and metabolic hormones showed smaller and more variable effects.

Due to the variability in study methodologies, measurement scales, and participant characteristics, the meta-analysis played a vital role in quantifying the overall effects. Although statistical heterogeneity measures such as I² and τ² were not computed, variability was visually represented in the forest plots. These plots highlighted the differences in effect sizes, particularly for secondary outcomes, with some studies showing larger effects and more modest results. The plots effectively summarise the findings and help illustrate areas of inconsistency, aiding the interpretation of the results.

All effect size conversions and calculations, including pooled SMDs and associated CIs, were conducted using Microsoft Excel (version 16.93.1), following established methodologies from the Cochrane Handbook^21^ and meta-analysis texts.^3,8^ No imputation methods were needed, as no missing data were identified, ensuring transparency and replicability.

In conclusion, the meta-analysis and forest plots provide a clear, quantitative summary of ISR effects. These visual and statistical representations offer insights into the magnitude of effects and variability across studies. Future research should address the variability in methodologies and participant characteristics to refine the understanding of optimal ISR for various training goals.

### Statistical Synthesis

Six studies were synthesised to assess the effects of inter-set rest intervals (<60s vs. >60s) on muscle hypertrophy, strength, metabolic hormones, power output, and motor unit recruitment. Standardised mean differences (SMDs) with 95% confidence intervals (CIs) were calculated to allow quantitative comparisons between the two ISR conditions, with pooled SMDs adjusted using Hedges’ g for small sample sizes.^3^

For muscle hypertrophy, the pooled SMD was 0.08 (95% CI: −0.28, 0.44), indicating a negligible difference between short and long rest intervals. Similarly, muscle strength showed a pooled SMD of −0.74 (95% CI: −1.19, −0.29), suggesting an advantage for longer ISR intervals. These findings were reflected in the forest plots (Figures 4, 5, and 6), which illustrate the pooled effect sizes and variability across studies.

For secondary outcomes, the pooled SMD for metabolic hormone responses was 0.11 (95% CI: −0.77 to 0.99), showing a small positive effect for longer ISR. Power output had a pooled SMD of −0.64 (95% CI: −1.2, −0.07), indicating a positive effect for longer ISR. Motor unit recruitment showed a pooled SMD of −0.66 (95% CI: −1.49, 0.16), suggesting a positive effect for longer ISR but with considerable uncertainty. Forest plots for these secondary outcomes were included in the final report to visually represent the variability in effect sizes.

Given the small number of included studies and the substantial variability in methodologies, populations, and outcome measures, formal heterogeneity statistics (e.g., I², τ²) were not calculated. In such contexts, these indices can be unstable and potentially misleading, as the variability is more plausibly attributable to systematic differences in study design and interventions rather than random error. Instead, heterogeneity was evaluated qualitatively through structured tabulation (Table 8) and visually using forest plots, which better illustrate the direction and magnitude of effects across studies. This approach provides a clearer representation of between-study variability while acknowledging the limitations of statistical heterogeneity measures in small, heterogeneous datasets.

**Table 8.**
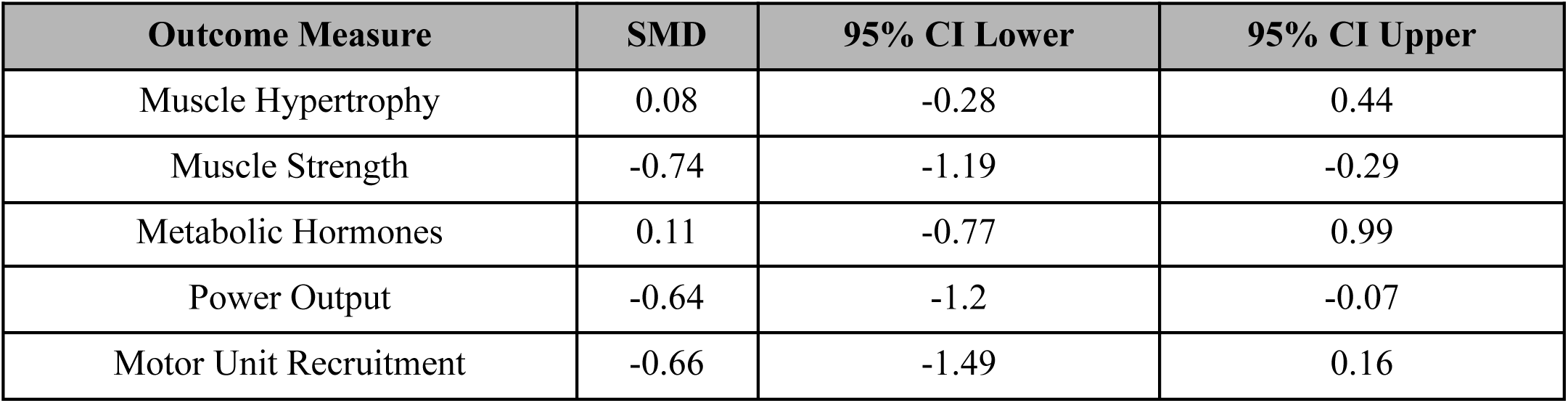
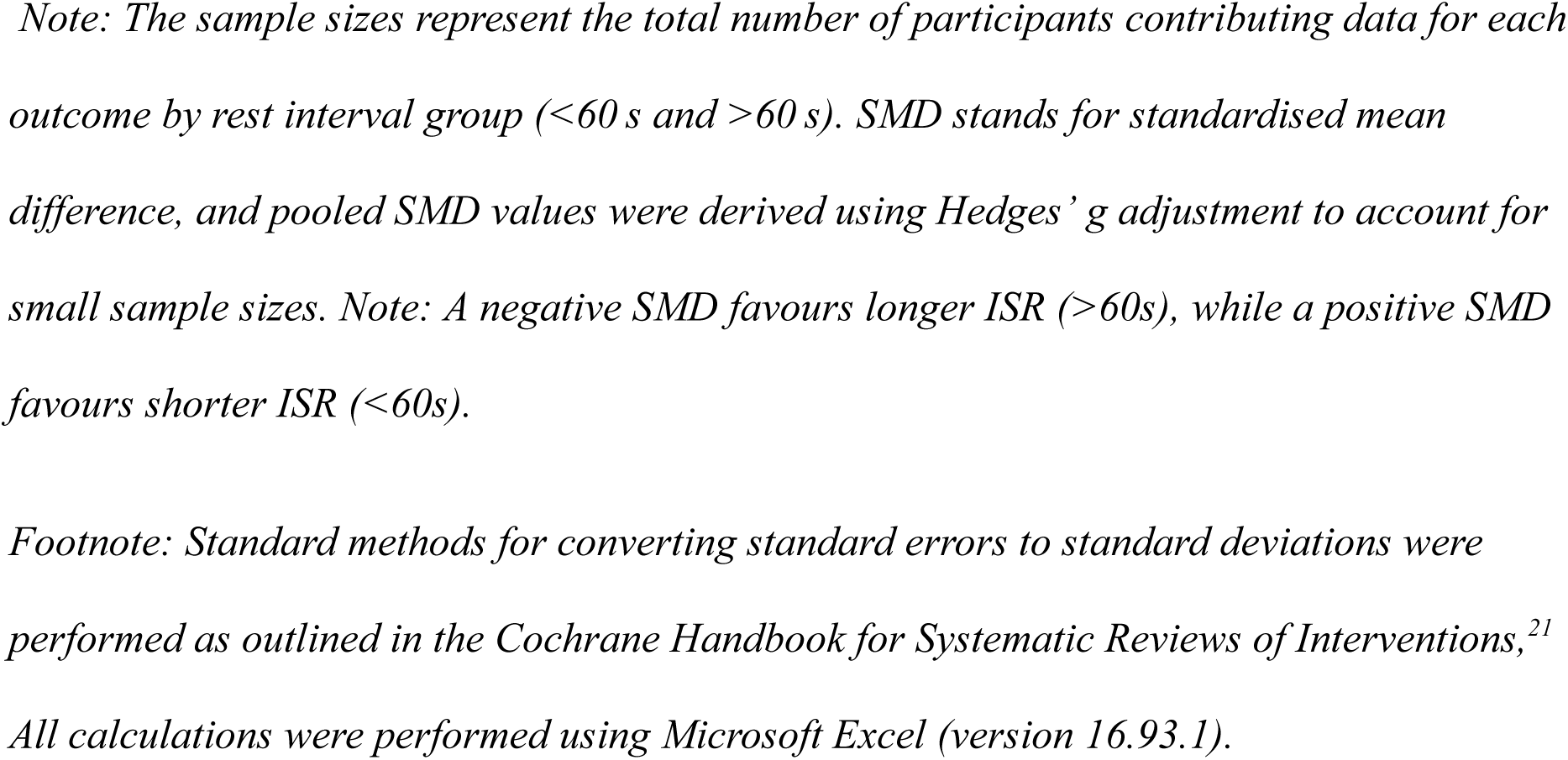
Statistical Synthesis of ISR Intervals on Key Outcomes Certainty Assessment.

| Outcome Measure | SMD | 95% CI Lower | 95% CI Upper |
| --- | --- | --- | --- |
| Muscle Hypertrophy | 0.08 | -0.28 | 0.44 |
| Muscle Strength | -0.74 | -1.19 | -0.29 |
| Metabolic Hormones | 0.11 | -0.77 | 0.99 |
| Power Output | -0.64 | -1.2 | -0.07 |
| Motor Unit Recruitment | -0.66 | -1.49 | 0.16 |

*Note: The sample sizes represent the total number of participants contributing data for each outcome by rest interval group (<60 s and >60 s). SMD stands for standardised mean difference, and pooled SMD values were derived using Hedges' g adjustment to account for small sample sizes. Note: A negative SMD favours longer ISR (>60s), while a positive SMD favours shorter ISR (<60s).*
*Footnote: Standard methods for converting standard errors to standard deviations were performed as outlined in the Cochrane Handbook for Systematic Reviews of Interventions,<sup>21</sup> All calculations were performed using Microsoft Excel (version 16.93.1).*
*Note: A negative SMD favours longer ISR (>60s), while a positive SMD favours shorter ISR (<60s).*

All calculations, including effect size conversions and data synthesis, were performed using Microsoft Excel (version 16.93.1) following standard systematic review methodologies.^21^ Detailed methods for converting effect estimates and calculating 95% CIs are provided in the supplementary material. These procedures were consistently applied to ensure transparency, replicability, and accuracy of the findings.

To visually synthesise the effects of inter-set rest intervals (ISR) on the various outcome measures, forest plots were generated for each primary outcome, including muscle hypertrophy (Figure 4), muscle strength (Figure 5), and the combined effect across all outcome measures (Figure 6). Forest plots are useful tools for summarising the overall effect estimates, visualising the variability between studies, and highlighting the direction and magnitude of the effects. Each plot represents the pooled standardised mean difference (SMD) for the specified outcome, with 95% confidence intervals (CIs) indicating the precision of the estimates. These plots facilitate a clear comparison of effect sizes across studies and provide insight into the consistency and heterogeneity of the results. The following sections present the forest plots for each outcome measure, offering a graphical overview of the impact of different ISR durations on hypertrophy, strength, and other secondary outcomes, supporting the synthesis of the findings discussed throughout the review.

**Figure 4.** Forest plot of hypertrophy outcomes comparing <60 s versus >60 s inter-set rest intervals across included studies. Standardised mean differences (SMD) with 95% confidence intervals are presented.

*Note: A negative SMD favours longer ISR (>60s), while a positive SMD favours shorter ISR (<60s)*.

This forest plot suggests that shorter inter-set rest intervals (<60 s) may lead to improvements in muscle hypertrophy. *Fink et al.* reported a large positive effect (SMD = 1.29), indicating a pronounced benefit of shorter rest intervals, whereas *Schoenfeld et al.* observed a small, non-significant positive effect (SMD = 0.11), suggesting only a trivial advantage. The variability in effect sizes likely reflects differences in study design, participant training status, and methodological approaches across studies.

**Figure 5.** Forest plot of strength outcomes comparing <60 s versus >60 s inter-set rest intervals across included studies. Standardised mean differences (SMD) with 95% confidence intervals are presented.

*Note: A negative SMD favours longer ISR (>60s), while a positive SMD favours shorter ISR (<60s)*.

This forest plot indicates considerable variability in the impact of inter-set rest intervals on muscle strength. *Fink et al.* reported a moderately positive effect (SMD = 0.65), suggesting a benefit of shorter rest intervals. By contrast, *Schoenfeld et al.* observed a small negative effect (SMD = −0.26), indicating a slight advantage for longer rest intervals. *De Salles et al.* reported a more pronounced negative effect (SMD = −1.31), further favouring longer rest intervals in their cohort. The divergence in results likely reflects methodological differences across studies, including variations in participant characteristics, training protocols, and outcome measures.

**Figure 6.** Forest plot of combined outcome measures (hypertrophy, power, strength, motor unit recruitment & metabolic hormones) across all included studies.

*Note: A negative SMD favours longer ISR (>60s), while a positive SMD favours shorter ISR (<60s)*.

This forest plot suggests that the effects of inter-set rest intervals on secondary outcomes vary considerably across studies. Hypertrophy (SMD = 0.08; 95% CI −0.28 to 0.44) and metabolic hormones (0.11; −0.77 to 0.99) showed negligible differences, with confidence intervals spanning zero. Motor unit recruitment (SMD = −0.66; 95% CI: −1.49 to 0.16) suggested a small, non-significant tendency favouring longer ISRs, though the wide confidence interval indicates considerable imprecision. In contrast, strength (SMD = −0.74; 95% CI −1.19 to −0.29) and power (−0.64; −1.20 to −0.07) showed statistically significant pooled effects favouring longer ISRs. Overall, these findings indicate that strength and power benefit modestly from longer ISRs, whereas hypertrophy, hormones, and motor unit recruitment remain inconsistent.

### Certainty of Evidence

The overall certainty of each outcome was assessed using the GRADE framework.^41^ Although all studies were randomised controlled trials, the certainty of specific outcomes was downgraded owing to study limitations, imprecision, and other concerns. Table 9 summarises the GRADE assessment for each outcome, including the number of studies, pooled effect estimates (SMDs with 95% CIs), overall GRADE ratings, and rationale for downgrading.

**Table 9.**
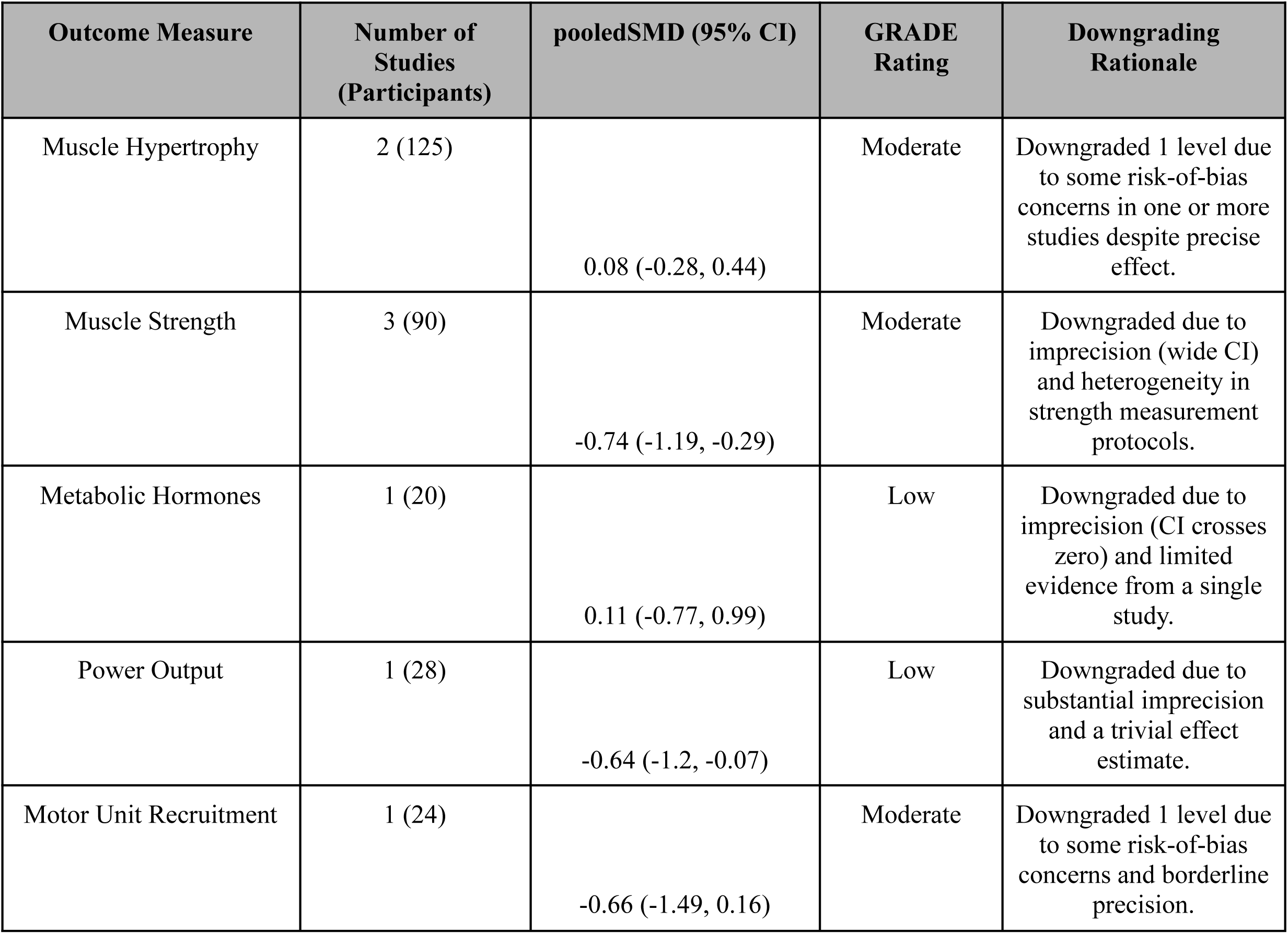
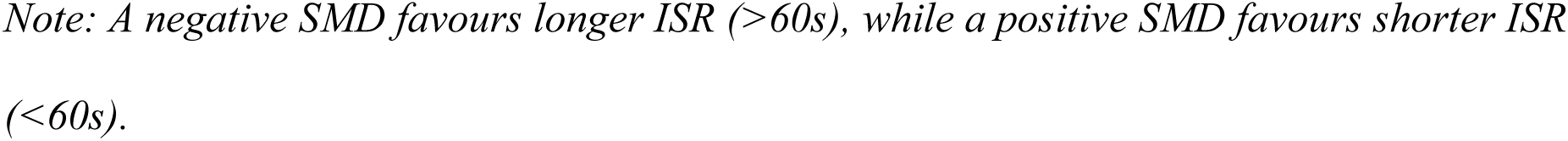
GRADE Summary of Findings for Key Outcomes.

| Outcome Measure | Number of Studies (Participants) | pooledSMD (95% CI) | GRADE Rating | Downgrading Rationale |
| --- | --- | --- | --- | --- |
| Muscle Hypertrophy | 2 (125) | 0.08 (-0.28, 0.44) | Moderate | Downgraded 1 level due to some risk-of-bias concerns in one or more studies despite precise effect. |
| Muscle Strength | 3 (90) | -0.74 (-1.19, -0.29) | Moderate | Downgraded due to imprecision (wide CI) and heterogeneity in strength measurement protocols. |
| Metabolic Hormones | 1 (20) | 0.11 (-0.77, 0.99) | Low | Downgraded due to imprecision (CI crosses zero) and limited evidence from a single study. |
| Power Output | 1 (28) | -0.64 (-1.2, -0.07) | Low | Downgraded due to substantial imprecision and a trivial effect estimate. |
| Motor Unit Recruitment | 1 (24) | -0.66 (-1.49, 0.16) | Moderate | Downgraded 1 level due to some risk-of-bias concerns and borderline precision. |

*Note: A negative SMD favours longer ISR (>60s), while a positive SMD favours shorter ISR (<60s).*

For muscle hypertrophy and strength, the evidence was rated as “moderate,” downgraded due to the risk of bias concerns and imprecision (wide CIs and variability in protocols). Evidence for metabolic hormones and power output was rated as “low” due to imprecision, small sample sizes, and trivial effect estimates. Motor unit recruitment was rated moderate, owing to minor risk of bias issues and borderline precision.

While formal tests for heterogeneity and publication bias were not conducted, a qualitative evaluation of these factors was used to guide downgrading decisions, ensuring the GRADE ratings accurately reflected the evidence’s strengths and limitations.

Table 9 summarises these findings and provides a transparent overview of the evidence regarding the effects of ISR intervals on muscle hypertrophy, muscle strength, metabolic hormone responses, power output, and motor unit recruitment.

## Discussion

Although caution is warranted in interpreting these findings owing to the small number of studies included, the current results appear to be largely in line with previous reviews on the effects of inter-set rest intervals (ISR) in resistance training. For example, recent reviews have noted that longer rest intervals (>60 s) are associated with enhanced strength and power. The present review demonstrated that long rest intervals (>60 s) are significantly associated with superior strength (SMD = −0.74, 95% CI: −1.19, −0.29) and power (SMD = −0.64, 95% CI: −1.2, −0.07) outcomes, suggesting that longer rest periods facilitate improved neuromuscular recovery and allow for heavier loads in subsequent sets. In contrast, the effects on muscle hypertrophy were minimal (SMD = 0.08, 95% CI: −0.28, 0.44), and the findings for metabolic hormone responses (SMD = 0.11, 95% CI: −0.77, 0.99) and motor unit recruitment (SMD = −0.66, 95% CI: −1.49, 0.16) were inconclusive, with confidence intervals that cross zero. These observations are consistent with studies indicating that, while longer ISR may slightly improve hypertrophic outcomes, the primary benefits of extended rest periods are realised in strength and power adaptations^12,22^. In line with this, the current review supports the recommendation that longer inter-set rest intervals should be employed when the primary goal is to maximise neuromuscular performance in resistance trained athletes, whereas shorter rest intervals may be more appropriate when the training objective is to induce metabolic stress, despite their apparent limitations in promoting maximal strength and power gains.

### Interpretation and Explanation of Findings

The positive effects of longer inter-set rest intervals (>60s) on hypertrophy and strength can be largely attributed to improved neuromuscular recovery, mechanical tension, and motor unit recruitment. Longer rest periods allow for more complete recovery of both the muscles and central nervous system (CNS), which reduces fatigue and enables individuals to lift heavier loads in subsequent sets. From a mechanistic stand-point, longer rest periods may enhance the ability to produce mechanical tension, which is vital for both hypertrophy and strength, potentially by reducing central neuromuscular fatigue, which aids in the production of internal fibre force that produces mechanical tension.^29^ The pooled standardised mean difference (SMD) for hypertrophy (0.08) and strength (−0.74) from the meta-analysis, although moderate to small, supports the benefit of longer rest intervals in promoting these outcomes. However, the variability in effect sizes across studies suggests that these effects may not always be significant, particularly in hypertrophy.

Shorter inter-set rest intervals (<60s) increase neuromuscular fatigue, limiting the amount of weight that can be lifted across sets.^6,10^ Inadequate recovery between sets can impede the full replenishment of energy stores and hinder the recruitment of high-threshold motor units, such as fast-twitch fibers, which are essential for maximal strength development. According to Henneman’s size principle, motor units are recruited in an orderly fashion from smallest to largest based on the force demands of the activity.^19^ During high-intensity resistance training, sufficient rest intervals (typically around 2 to 3 minutes) are crucial to allow for the resynthesis of adenosine triphosphate (ATP) and phosphocreatine (PCr), which are rapidly depleted during intense efforts. This recovery enables the nervous system to effectively activate larger, high-threshold motor units necessary for generating maximal force in subsequent sets. Studies have demonstrated that longer rest periods facilitate greater motor unit recruitment and improved performance in resistance training sessions^2,7^. As fatigue accumulates, force production decreases, hindering optimal hypertrophy and strength outcomes.^4^ This is supported by the relatively small or negligible effect sizes observed for power output (SMD = −0.64, 95% CI: −1.2, −0.07) and metabolic hormones (SMD = 0.11, 95% CI: −0.77, 0.99), underscoring the limited impact of shorter ISR on these secondary outcomes.

The importance of mechanical tension in muscle growth was further supported by these results. A longer ISR allows for the lifting of heavier loads, thus supporting progressive overload, a critical factor in hypertrophy and strength development. In contrast, a shorter ISR limits the weight lifted per set, reducing the overall mechanical tension applied to the muscle and hindering hypertrophy and strength adaptations.

Motor unit recruitment is another factor that is influenced by ISR duration. A longer ISR allows for the recruitment of high-threshold motor units, including fast-twitch fibres, which are essential for maximal strength and hypertrophy.^31^ According to Henneman’s Size Principle (1957), extended rest periods ensure that these motor units are recruited across all sets, thus providing a greater hypertrophic stimulus. In contrast, shorter rest intervals reduce the recruitment of high-threshold motor units, thereby diminishing strength potential.

Although a longer ISR is mainly associated with increased mechanical tension and motor unit recruitment, a shorter ISR can lead to greater metabolic stress, which promotes hypertrophy through mechanisms such as cell swelling, metabolite accumulation, and hormonal responses.^38^ However, although metabolic stress may aid hypertrophy, it does not enhance strength development, which relies on the ability to lift maximal loads. Therefore, while a shorter ISR may promote hypertrophy through metabolic stress, it is less effective for strength gains because of its inability to facilitate maximal force production.

### Limitations of the Evidence

Although the review indicated moderate effects of inter-set rest intervals (ISR) on hypertrophy and strength, several limitations must be acknowledged. The study populations were predominantly resistance-trained males, and the number of studies reporting secondary outcomes, particularly power output and metabolic hormones, was relatively small, thereby limiting the precision of these effect estimates. The included studies were frequently constrained by small sample sizes, varied ISR protocols, and considerable heterogeneity in exercise designs and participant characteristics, all of which challenged the generalisability of the findings. Furthermore, inconsistencies in the reporting of methodological details, such as exposure duration and protocol standardisation, have hindered the ability to draw definitive conclusions regarding the impact of ISR on these outcomes.

### Limitations of Review Processes

The data extraction process was conducted by a single reviewer using a pre-designed form tailored to the objectives of the review, and the extracted data were verified by a second reviewer to ensure accuracy and resolve any discrepancies. While this process adhered to the approach recommended by Cochrane for systematic reviews, where single data extraction with verification can be valid when double extraction is not feasible owing to time or resource constraints,^21,35^ it introduces potential errors, particularly when discrepancies between reviewers can not be resolved swiftly. Furthermore, owing to the varied reporting methods and incomplete data in some studies, assumptions were not made. Instead, attempts were made to contact the authors for clarification and additional details. However, some missing information may have still affected the final analysis.

Although structured workflows and predefined criteria were employed to ensure consistency in data extraction and quality assessment, reliance on a single reviewer for the initial data extraction step increases the risk of potential biases. These biases were mitigated by a second reviewer verifying the data; however the lack of independent data extraction by the two reviewers for all studies remains a limitation. Additionally, given the wide variability in reporting across studies, some outcome measures were not directly comparable, necessitating conversions and imputation of missing data. While these transformations were applied consistently across studies and are detailed in the methods section, there remains a possibility that some inaccuracies or inconsistencies could have influenced the results.

The review also included only English-language studies, which could have led to the exclusion of valuable data published in other languages. Despite these limitations, the structured and transparent approach to the review process minimised potential biases, and the methods employed were not anticipated to substantially alter the overall conclusions regarding the impact of ISR on hypertrophy and strength outcomes.

### Practical Implications for Training

The findings from the review suggest several practical recommendations for resistance training, particularly when prescribing inter-set rest intervals (ISR) for resistance-trained athletes. For individuals focused on improving both muscle hypertrophy and strength, Longer rest intervals (>60s) are likely to be more effective and may warrant prioritisation in strength-focused programmes. These longer intervals facilitate better recovery between sets, allowing for the use of heavier loads, which is crucial for both strength development and maximising mechanical tension. The ability to lift progressively heavier weights in subsequent sets enhances the stimulus for both strength and hypertrophic adaptation.

While shorter rest intervals (<60s) can still be beneficial, particularly for inducing metabolic stress, which is thought to contribute to hypertrophy through mechanisms such as cell swelling and hormonal responses, they may not be as effective in maximising both strength and hypertrophy outcomes in resistance-trained individuals. Given the minimal effect on hypertrophy observed in the review (SMD = 0.08, 95% CI: −0.28, 0.44), and the significant strength benefits provided by longer rest intervals (SMD = −0.74, 95% CI: −1.19, −0.29). Current evidence suggests that longer ISR may be more beneficial for those seeking optimal results in strength development.

Therefore, for athletes and resistance trainees looking to enhance strength performance, prioritising longer rest intervals (>60s) is likely to provide superior outcomes. For individuals specifically aiming for hypertrophy, shorter intervals may still have some benefit, but the benefits may not be as pronounced as once thought, particularly when maximal load lifting is a key goal. Thus, incorporating a longer ISR for strength-focused training and a shorter ISR for hypertrophy-focused metabolic stress training might offer a balanced approach depending on the primary training goal.

### Recommendations for Future Research

Future research should focus on the variability in inter-set rest (ISR) responses, particularly with regard to individual factors such as training experience, sex, and genetic predispositions. The current body of literature predominantly includes resistance-trained males, with limited representation from other populations, such as females or individuals with different training backgrounds. Investigating how these demographic and genetic factors influence ISR responses would provide valuable insights for optimising training protocols tailored to individual needs.

Moreover, additional longitudinal studies are required to assess the long-term effects of ISR duration on hypertrophy, strength, and other related outcomes. Although short-term studies provide useful insights, understanding the sustained impact of different ISR durations over extended periods is essential for developing comprehensive training regimens.

Standardising ISR protocols across studies, especially concerning outcome measures and rest interval durations, would greatly enhance the comparability of results and improve the consistency of conclusions drawn from literature. This standardisation would enable more robust meta-analyses and provide clearer guidance to practitioners in the field.

In addition, future studies should investigate the broader, long-term impacts of ISR variations on performance outcomes, including athletic performance, injury prevention, and overall training adaptation. Exploring these areas will allow researchers to better understand the multifaceted effects of ISR on physical performance, further informing training strategies across various populations and training objectives.

## Conclusion

In summary, the review highlights the positive effects of longer inter-set rest intervals (>60s) on muscle hypertrophy and strength, which may reflect more complete recovery between sets, enabling greater force production, thereby enhancing the ability to lift heavier loads and promoting muscle growth. The findings align with existing literature suggesting that longer rest intervals may be beneficial for maximal strength and hypertrophy adaptations, as they facilitate sustained mechanical tension and optimal motor unit recruitment across sets. In contrast, shorter rest intervals are associated with increased metabolic demand, which has been hypothesised to contribute to hypertrophy,^38^ However, the benefits of shorter rest intervals on strength and hypertrophy appear less pronounced than those of longer rest intervals.

Future research should continue to explore how inter-set rest intervals can be optimised for different training goals and populations. This includes investigating factors such as age, sex, training experience, and individual variability in response to the rest duration. Longitudinal studies are needed to better understand the long-term impacts of different ISR strategies on performance, injury prevention, and overall training adaptation. Ultimately, refining the current understanding of ISR will enable more tailored and effective training regimens, maximising outcomes for athletes and recreational lifters alike.

## Registration and Protocol

The systematic review was registered in the International Prospective Register of Systematic Reviews, PROSPERO, under registration number: CRD42024590003. Registration entry outlines the original protocol for the review, including predefined inclusion criteria, outcome measures, and statistical methods. The registered protocol was followed, with no major deviations. Any modifications to the protocol made during the review process were clearly documented and justified in the final report, ensuring transparency and minimising potential bias in the systematic review process.

## Support

The systematic review received no specific financial or non-financial support for its execution. The review was conducted independently without external funding or sponsorship. As such, no funders played a role in the design of the review, data collection, analysis, decision to publish, or preparation of the manuscript. All research activities, including study selection, data synthesis, and manuscript preparation, were performed by the authors. Therefore, there is no potential for bias arising from external involvement during the review process.

## Contributorship

LD conceived and designed the review, conducted data extraction, and drafted the manuscript. SB independently verified data extraction, contributed to analysis, and critically revised the manuscript. LD is the guarantor.

## Supporting information

Figure 2. Traffic-light plots of domain-level risk-of-bias judgements for each individual result.

Figure 5. Forest plot of the effects of inter-set rest intervals on muscle strength across included studies.

Figure 4. Forest plot of the effects of inter-set rest intervals on muscle hypertrophy across included studies.

Figure 1. PRISMA 2020 flow diagram for systematic reviews based on database searches.

Figure 6. Forest plot of the combined effects of inter-set rest intervals on hypertrophy, strength, motor unit recruitment, hormonal outcomes, and pow

Figure 3. Weighted bar plots showing the distribution of risk-of-bias judgements across bias domains.

## Data Availability

All data produced in the present study are available upon reasonable request to the authors

## Acknowledgements

None.

## Ethical approval

Not applicable (systematic review and meta-analysis of published studies).

## Availability of Data, Code, and Other Materials

The data, codes, and materials relevant to the systematic review will be made available upon request. This included the data collection forms, data extracted from the included studies, and the analytic code used in the meta-analysis. Interested researchers can contact the corresponding author to request access to these materials. We are committed to transparency and will provide the necessary data and code for the replication of these findings and further research, subject to legal or ethical restrictions.

