## Supplementary figures and images for "Investigating the impact of less than or greater than 60 seconds of inter-set rest on muscle hypertrophy and strength increases in males with >1 year of resistance training experience: systematic review with meta-analysis"

### Figure 1. PRISMA 2020 flow diagram for systematic reviews based on database searches.

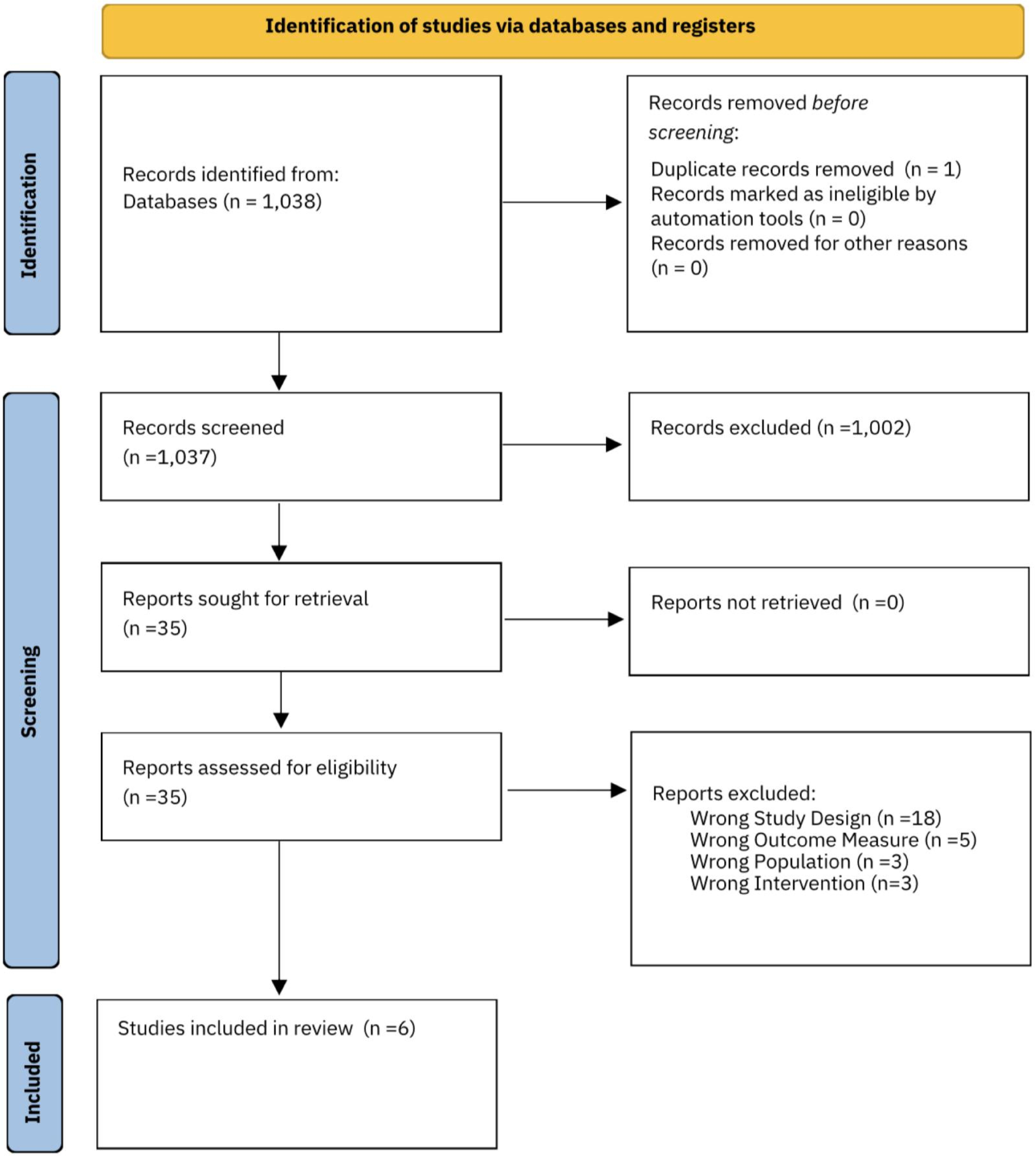

### Figure 2. Traffic-light plots of domain-level risk-of-bias judgements for each individual result.

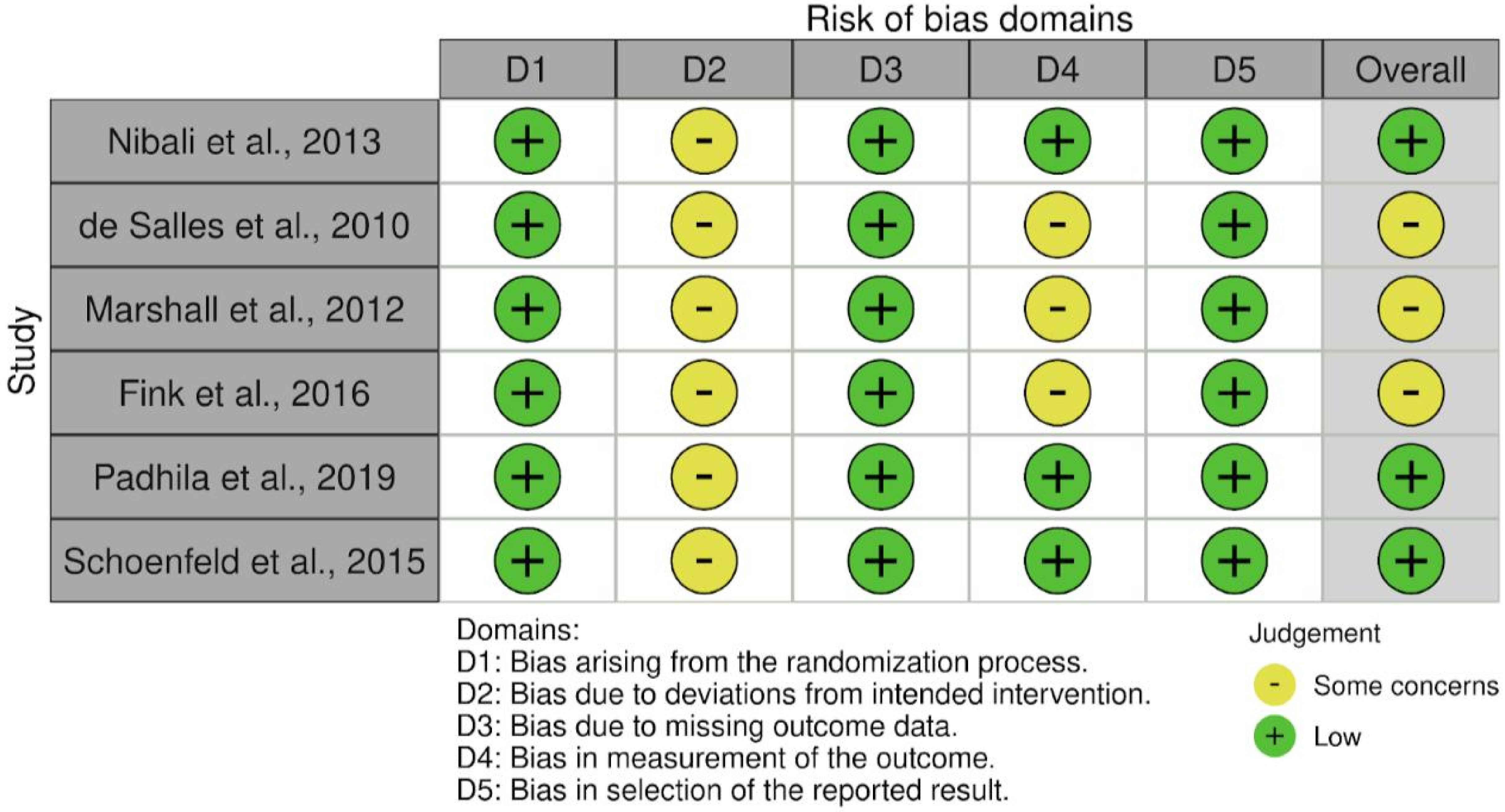

### Figure 3. Weighted bar plots showing the distribution of risk-of-bias judgements across bias domains.

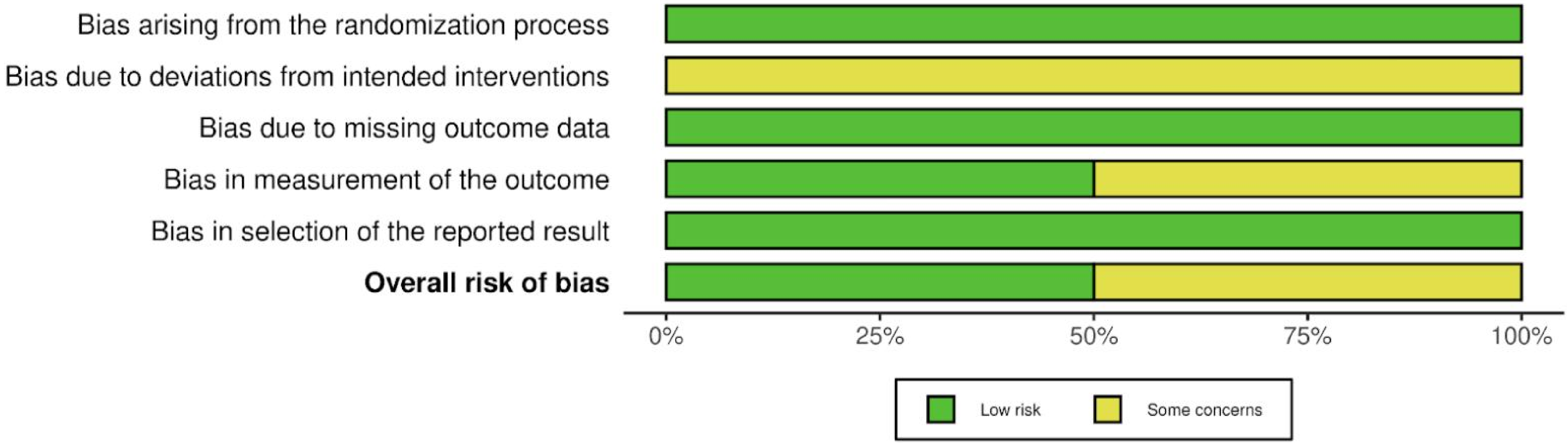

### Figure 4. Forest plot of the effects of inter-set rest intervals on muscle hypertrophy across included studies.

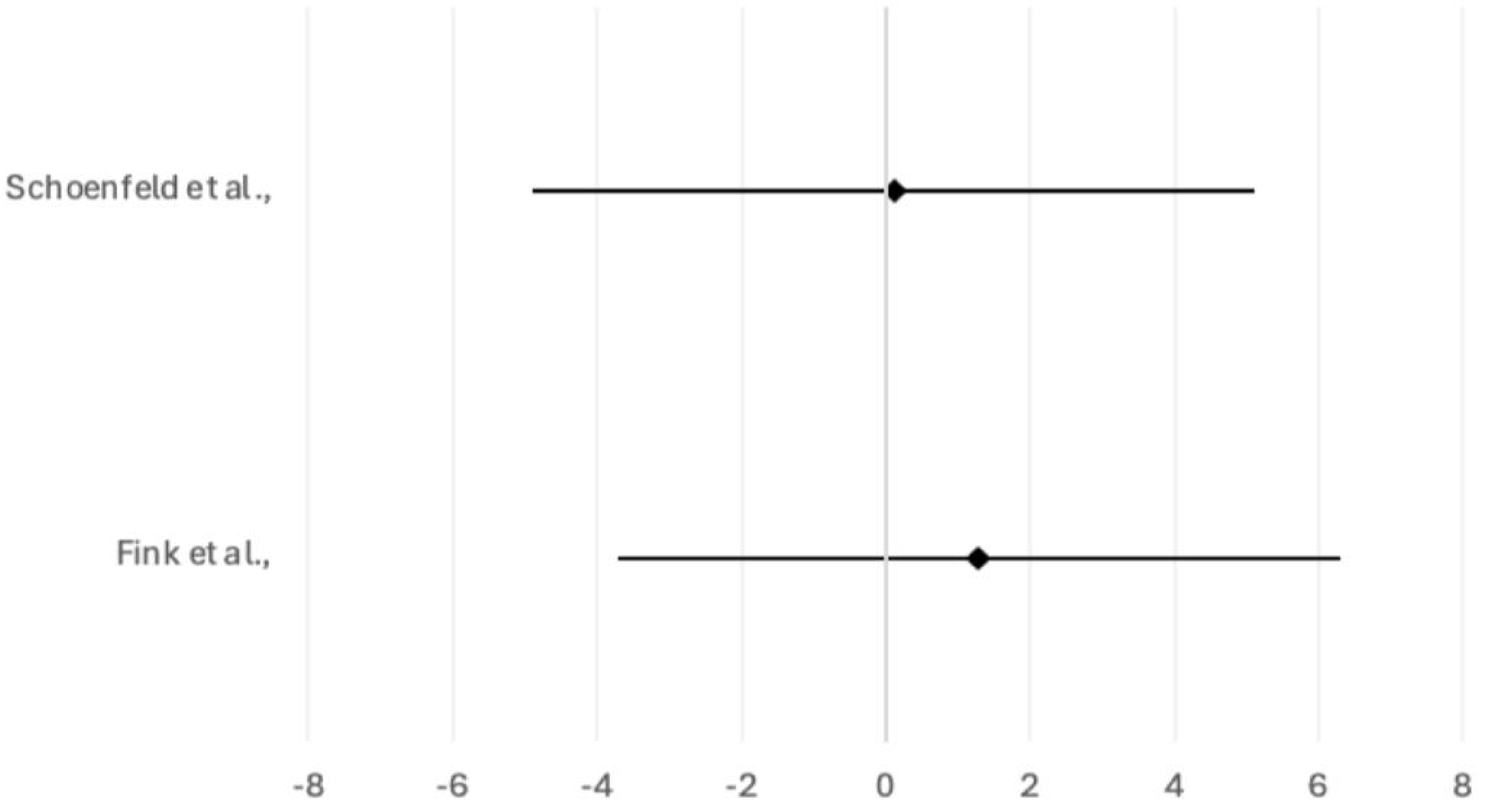

### Figure 5. Forest plot of the effects of inter-set rest intervals on muscle strength across included studies.

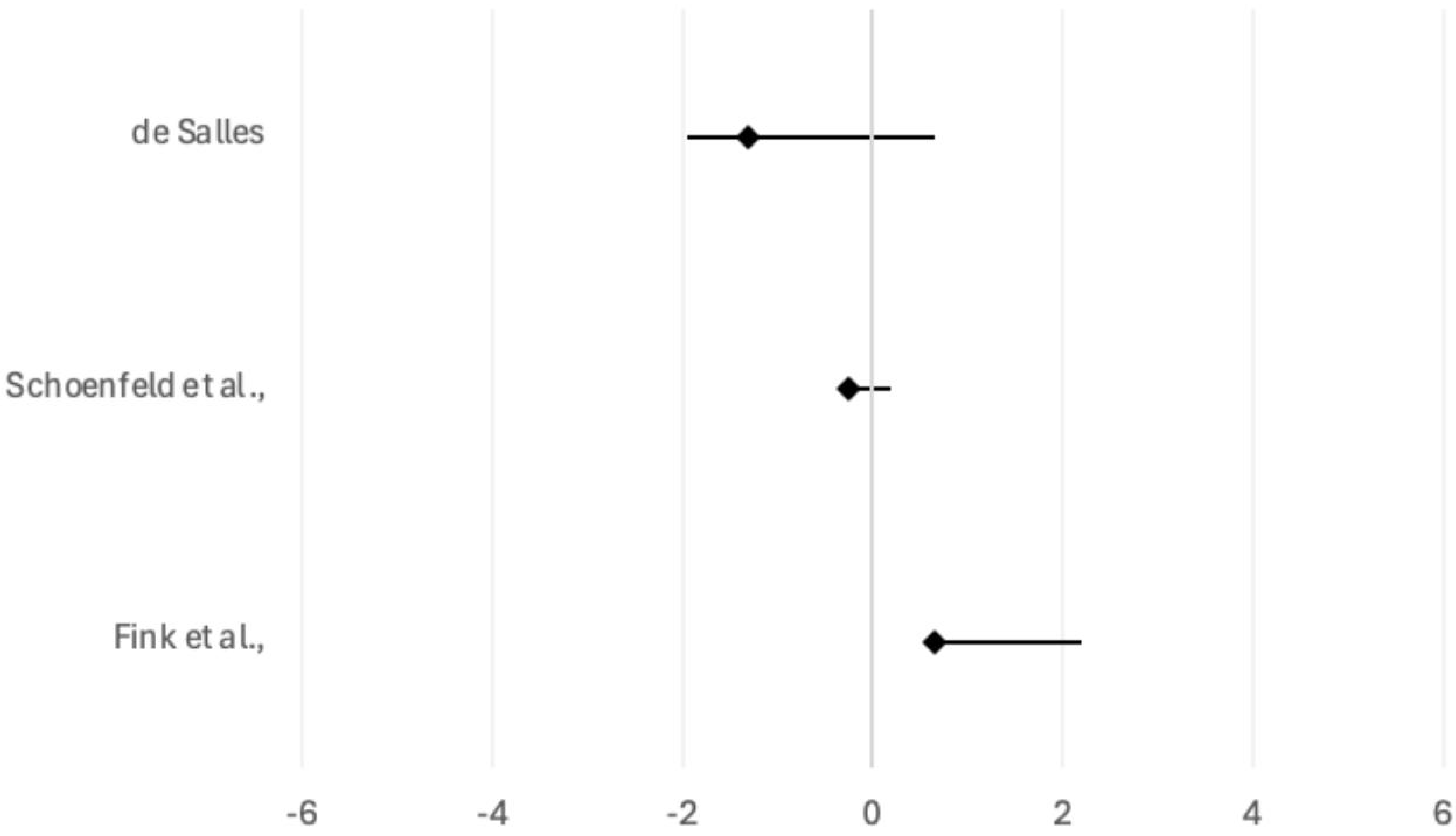

### Figure 6. Forest plot of the combined effects of inter-set rest intervals on hypertrophy, strength, motor unit recruitment, hormonal outcomes, and pow

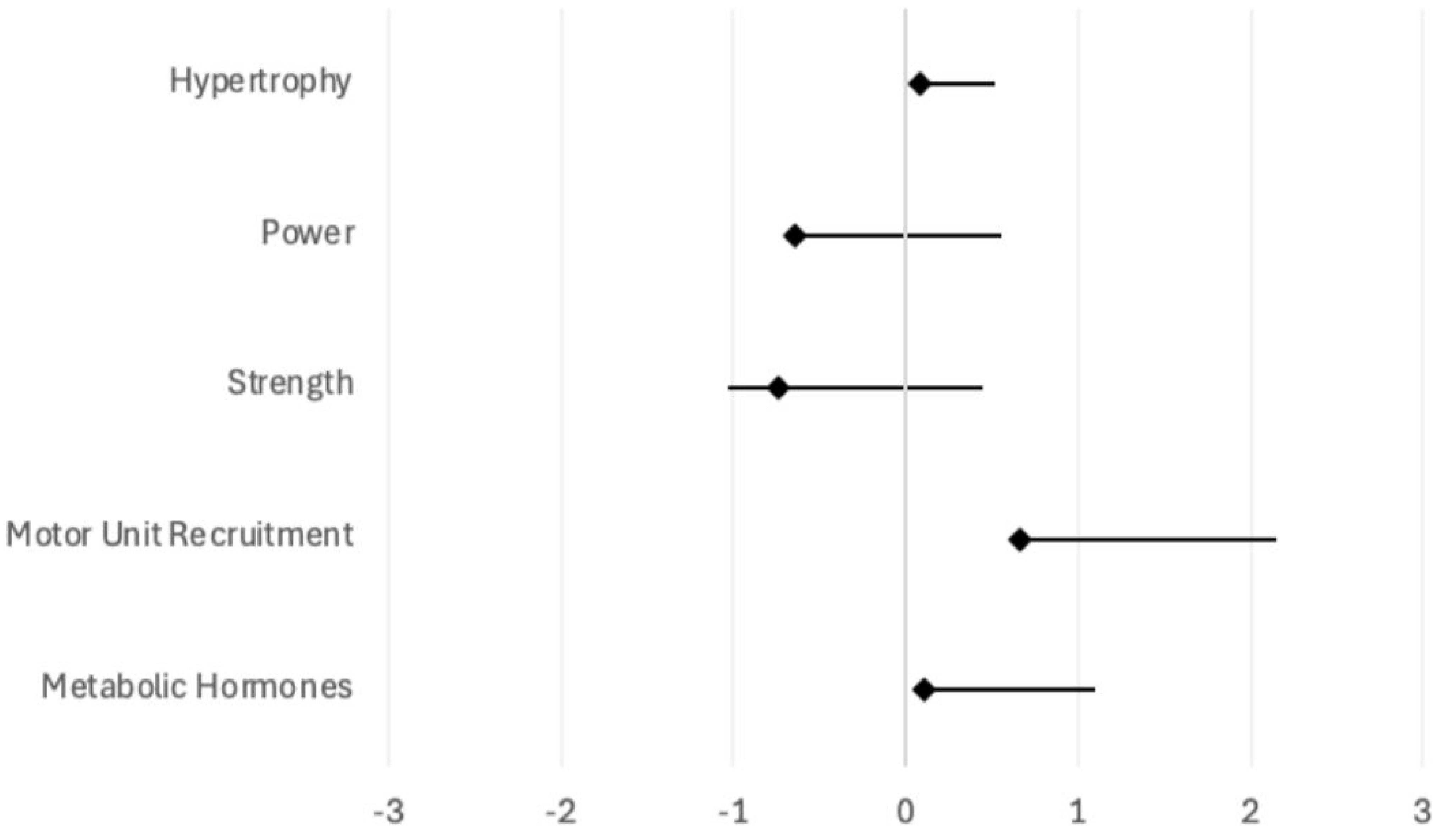
